# Forecasting invasive mosquito abundance in the Basque Country, Spain using machine learning techniques

**DOI:** 10.1101/2025.01.02.25319885

**Authors:** Vanessa Steindorf, K. B. Hamna Mariyam, Nico Stollenwerk, Aitor Cevidanes, Jesús F. Barandika, Patricia Vazquez, Ana L. García-Pérez, Maíra Aguiar

## Abstract

Mosquito-borne diseases cause millions of deaths each year and are increasingly spreading from tropical and subtropical regions into temperate zones, creating significant public health risks. The establishment of mosquito species in new areas increases the risk of local transmission (autochthonous cases), driven by both rising mosquito populations and viremic imported cases, infected travelers who can spark local transmission. Such developments present new challenges for public health systems in non-endemic regions.

In Spain, in the Basque Country region, the spread of mosquitoes, driven by changing climatic conditions, has enhanced mosquito adaptation alongside an increase in imported cases of dengue, Zika, and chikungunya. By employing a model that captures the complexities of the mosquito life cycle driven by the interaction with weather variables, including temperature, precipitation, and humidity, and leveraging machine learning techniques, this study aims to predict *Aedes* invasive mosquito abundance in provinces of the Basque Country, using egg count as a proxy and the weather features as key independent variables.

Statistical analyses explored the impact of temperature, precipitation, and humidity on mosquito egg abundance. Using lagged climate variables and ovitrap egg counts, models were evaluated using Root Mean Squared Error (RMSE) and Mean Absolute Error (MAE) metrics. The Random Forest (RF) model demonstrated the highest accuracy, followed by the Seasonal Autoregressive Integrated Moving Average (SARIMAX) model. Lastly, the best models were implemented to forecast *Aedes* invasive mosquito abundance in the Basque Country provinces. This forecasting tool aids vector control strategies in regions with expanding mosquito populations, highlighting the need for ongoing entomological surveillance to improve mosquito spread assessments.

## 1 Introduction

Vector-borne diseases, particularly those transmitted by mosquitoes, have become a significant global concern. The expansion of mosquitoes and the increase in transmitted diseases are escalating worldwide [18]. In the Americas, dengue cases alone surpassed 7 million by May 2024, exceeding the total annual of 4.6 million cases reported in the previous year [44]. Traditionally, these diseases primarily affected tropical and subtropical regions [18, 21, 44]. However, climate change and global warming are facilitating the spread, adaptation, and establishment of competent mosquitoes into temperate zones previously unaffected by such diseases, such as Europe [11]. Additionally, increased human mobility also plays a critical role, as travelers returning from endemic areas to non-endemic regions may introduce infections (imported cases), potentially sparking local transmission in areas with competent vectors and susceptible populations. Recently, countries like France, Italy, and Spain have experienced a significant rise in dengue imported cases. In France, from the beginning of 2024 up to June of the same year, the imported cases overpasses the 200 cases recorded over the whole previously year (in 2023) [35]. And, around 500 imported cases were registered in Italy. Additionally, there has been a marked increase in autochthonous cases, with 85 reported in France and 207 in Italy [18].

In the Basque Country, an autonomous community in northern Spain, no autochthonous cases of *Aedes* mosquito-borne diseases have been recorded to date. However, with the lifting of mobility restrictions after the SARS-CoV-2 pandemic, the Public Health Epidemiological Unit in the Basque Country has registered an increase of dengue, chikungunya, and Zika imported cases [32]. On the other hand, entomological surveillance in various localities has shown an increase in the abundance of *Aedes albopictus* eggs, and the establishment of *Aedes japonicus* populations [9]. These developments highlight the critical importance of maintaining robust surveillance systems, as effective monitoring is essential for preventing and controlling the spread of arboviruses.

The mosquitoes undergo to three life stages before becoming adults: egg, larva, and pupa. Female mosquitoes search for human blood since it provides the essential nutrients required for egg development. After feeding, the female typically rests while her eggs mature, and then lays them in small batches in areas with stagnant water, such as containers, tire ruts, or tree holes. A female mosquito can lay an average of 200 to 400 eggs at a time [17, 31]. Most eggs hatch into larvae within 48 hours if still water is available. However, they can survive several days, from 300 to 400 days, without coming into contact with water [17, 31]. This reproductive process is strongly influenced by environmental factors such as temperature, humidity, and rainfall, which affect the availability of suitable breeding sites and ultimately the success of egg development.

The worst conditions for *Aedes albopictus* eggs are high temperatures and low relative humidity [25]. Egg mortality decreases with increasing relative humidity and median temperatures of 24-26°*C*. Conversely, the optimum temperature for females to lay eggs is between 25-30°*C*. At temperatures of 20°*C* and 34°*C*, mosquitoes lay significantly fewer eggs [20, 25]. The optimal temperature for the development and survival of *Aedes albopictus* occurs at summer temperatures of 25-30°*C*. While a mean winter temperature of more than 0°*C* allows egg survival, a mean annual temperature of more than 11°*C* is required for adult activity [18]. At least 500 mm of annual rainfall is required for the breeding habitat, although mosquito populations have been established in areas with lower rainfall [1]. In contrast, periods of high precipitation temporarily reduce the number of females actively searching for a host. The reproductive season is influenced by increasing temperatures in spring and the onset of egg diapause in autumn, triggered by daylight hours below 13-14 hours [1, 18].

The association between climate factors and the prediction of dengue outbreaks has been widely studied [5, 7, 10, 12, 14, 26, 33]. By employing machine learning approaches, particularly those applied in endemic regions, have shown promise in enhancing the accuracy of dengue outbreak forecasts. On the other hand, some studies have incorporated vector data, such as adult mosquito populations, as proxies [12, 16, 27, 41], or larvae abundance [40]. Moreover, the role of *Aedes aegypti* abundance, climatic factors, and disease surveillance has been also evaluated in regions where autochthonous dengue transmission was recently introduced, such as in southern Brazil [12].

However, one of the key challenges in fitting and validating predictive models is the necessity of local incidence data on mosquito-borne diseases cases and vector surveillance information. This data serves as a critical predictor variable for outbreak forecasting, but it is typically only available in endemic regions, where autocthonous cases is a persistent public health concern. Unfortunately, such data is often limited or spatially restricted due to various factors, primarily the high costs associated with collecting and maintaining accurate, up-to-date surveillance systems, making it difficult to obtain comprehensive data for non-endemic or under-resourced regions [15].

Despite these challenges, numerous studies have successfully used climate variables and also historical data on mosquito adult abundance as proxies to forecast mosquito abundance. For example, mosquito abundance has been predicted using artificial neural network (ANN) models [27, 28], with some studies using adult mosquito populations as predictors [27], while others employed mechanistic models [38]. Another study used an ordinary differential equation (ODE) model to predict mosquito abundance, considering temperature, rainfall, egg diapause, and population dynamics of mosquitoes in southern France [43]. Nonetheless, this study did not include humidity as a climate factor, and prior hypotheses based on vector-related parameters were necessary, drawn from existing literature.

Finally, only a few studies have considered mosquito eggs as predictors for temporal forecasting [5, 8, 10]. A more recent study employed spatio-temporal forecasting using stacked machine learning techniques [13]. Most studies that have used egg counts for forecasting have linked them with climate changes and ovitrap data to predict dengue outbreaks in endemic regions. However, in non-endemic areas like the Basque Country, where there is no local *Aedes* mosquito-borne diseases transmission and adult mosquito populations are not systematically monitored, predicting mosquito abundance becomes crucial for controlling the spread of the disease and informing surveillance and intervention strategies.

In this study, we aim to estimate *Aedes* invasive mosquito abundance in a region where autochthonous mosquito-borne diseases transmitted by *Aedes albopictus* (such as dengue) have not yet been recorded, such as the Basque Country. By using the available data from the Basque Country’s provinces, we use machine learning techniques to model the relationship between recorded mosquito ovitrap egg counts and key environmental factors, including temperature, humidity, and precipitation. In Section 2, the relationship between climate variables and the abundance of mosquito eggs is analyzed within the context of a maritime climate, as the Basque Country, at the provincial and municipality levels. We explore and compare different machine learning models, considering variations such as including and excluding lagged versions of egg counts as a predictor, in Section 3. Notably, incorporating lagged versions of both independent and dependent variables consistently improves the performance of most models, demonstrating the importance of temporal dependencies in mosquito abundance forecasting. Further, in Section 3, fitting the best-performing models to the available data on recorded egg counts in ovitraps allow us to produce more accurate predictions of invasive mosquito abundance.

## 2 Materials and Methods

### 2.1 Entomological and meteorological data

Data on *Aedes* mosquito egg counts from 2013 to 2023 in the Basque Country were obtained using ovitraps as described in [9, 22]. Following the European Centre for Disease Prevention and Control (ECDC) recommended guidelines [18], the ovitraps were distributed across the three provinces, covering 63 municipalities, as shown in Figure 1(b).

**Figure 1:**
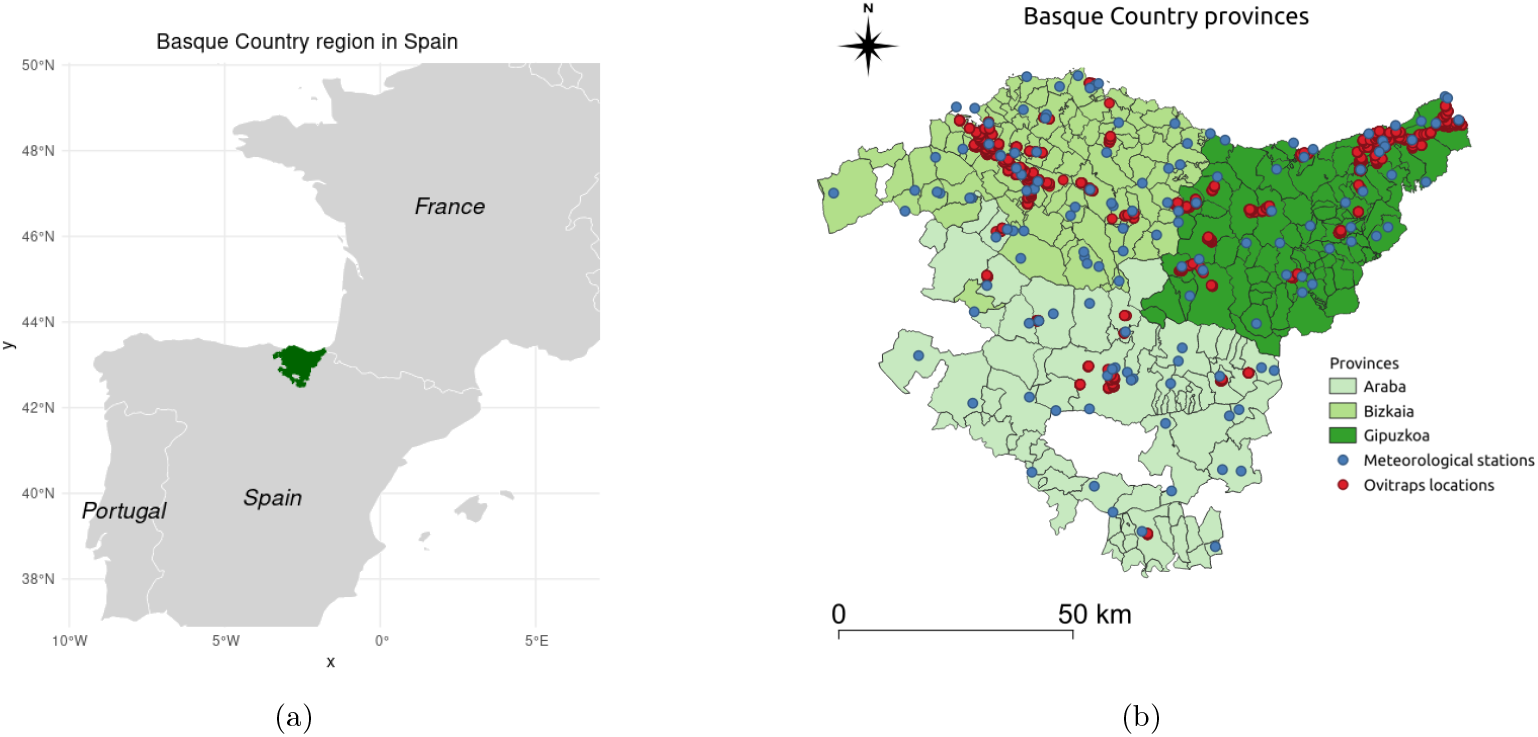
(a) Basque Country region in Spain (location in the European map). (b) Meteorological stations and ovitraps locations in the Basque Country provinces during the intersection study period (2016 to 2023).

The number of ovitraps varies by municipality, with two sampling areas selected in most cases. Each sampling area typically contains five ovitraps, which are positioned in sheltered spots away from direct sunlight and wind, often hidden within vegetation. Therefore, up to 10 ovitraps per municipality were placed in most cases. Each ovitrap contains water and a wooden stick (or tablex) that serves as a substrate for mosquito egg-laying. Every 14 days (on average), these paddles are removed, and new ones are put in their place. Thus, each municipality and area is sampled roughly 10 to 12 times per year, from June through November [9].

Meteorological data for the Basque Country were collected from the Basque Meteorological Agency (Euskalmet) across several weather stations (see Figure 1(b)) ^1^, covering the period from 2016 to 2023. The data, obtained from the OpenData Euskadi website [29], include precipitation (recorded as cumulative precipitation in millimeters (*mm*) or liters per square meter (*l/m*^2^)), temperature (measured in degrees Celsius (°*C*)), and humidity (relative air humidity as a percentage (%)). Weather observations were recorded every 10 minutes at each station. For this study, we calculated daily averages of temperature and humidity and daily cumulative precipitation for each meteorological station.

#### 2.1.1 Study area and data per provinces

The Basque Country, located in northern Spain, is divided into three administrative provinces: Araba (Álava), Bizkaia (Biscay), and Gipuzkoa (see Figure 1). With a total area of 7234 *km*^2^ and a population of approximately 2.18 million [19], the region is characterized by diverse landscapes and a maritime climate, with temperate conditions and high annual precipitation, particularly in the coastal areas. Araba, the southernmost province, has a more continental influence in its climate, with drier and slightly colder conditions than the coastal provinces of Bizkaia and Gipuzkoa. Bizkaia and Gipuzkoa, bordered by the Cantabrian Sea, experience milder temperatures and higher humidity. These climatic differences across the provinces influence the mosquito abundance patterns, which this study aims to capture and analyze through the environmental data collected.

For this study, we analyzed ovitrap mosquito egg counts collected in various locations across all three provinces. The data was pre-processed by averaging the 20 highest egg counts per province over a 14-day interval, considering that each municipality had a maximum of 10 ovitraps. This approach was necessary to address inconsistencies in the number of monitored ovitraps over the studied period and to avoid skewing the results with prevalent zero counts. By selecting the 20 largest egg counts, the data reflects meaningful mosquito activity (in at least two distinct locations), effectively filtering out areas with consistently low or zero activity.

Meteorological data, specifically daily precipitation (cumulative precipitation in millimeters (*mm*), air temperature (in degrees Celsius (°*C*)), and relative humidity (percentage (%)), were obtained by averaging daily values from all available meteorological stations in each province. These features were then aggregated over the previous 14 days to maintain consistent time intervals between the entomological and meteorological datasets. The average annual temperature and accumulated precipitation in each province align with environmental conditions favorable for *Aedes albopictus* survival, approximately 11.5 °*C* and 878 *mm* in Araba, 13.8 °*C* and 1278 *mm* in Bizkaia, and 13.4 °*C* and 1610 *mm* in Gipuzkoa [9], which are consistent with the survival thresholds discussed in the literature for this species [1, 18].

The time series of the average egg counts, temperature, humidity, and cumulative precipitation for each province in the Basque Country are shown in Figure 2.

**Figure 2:**
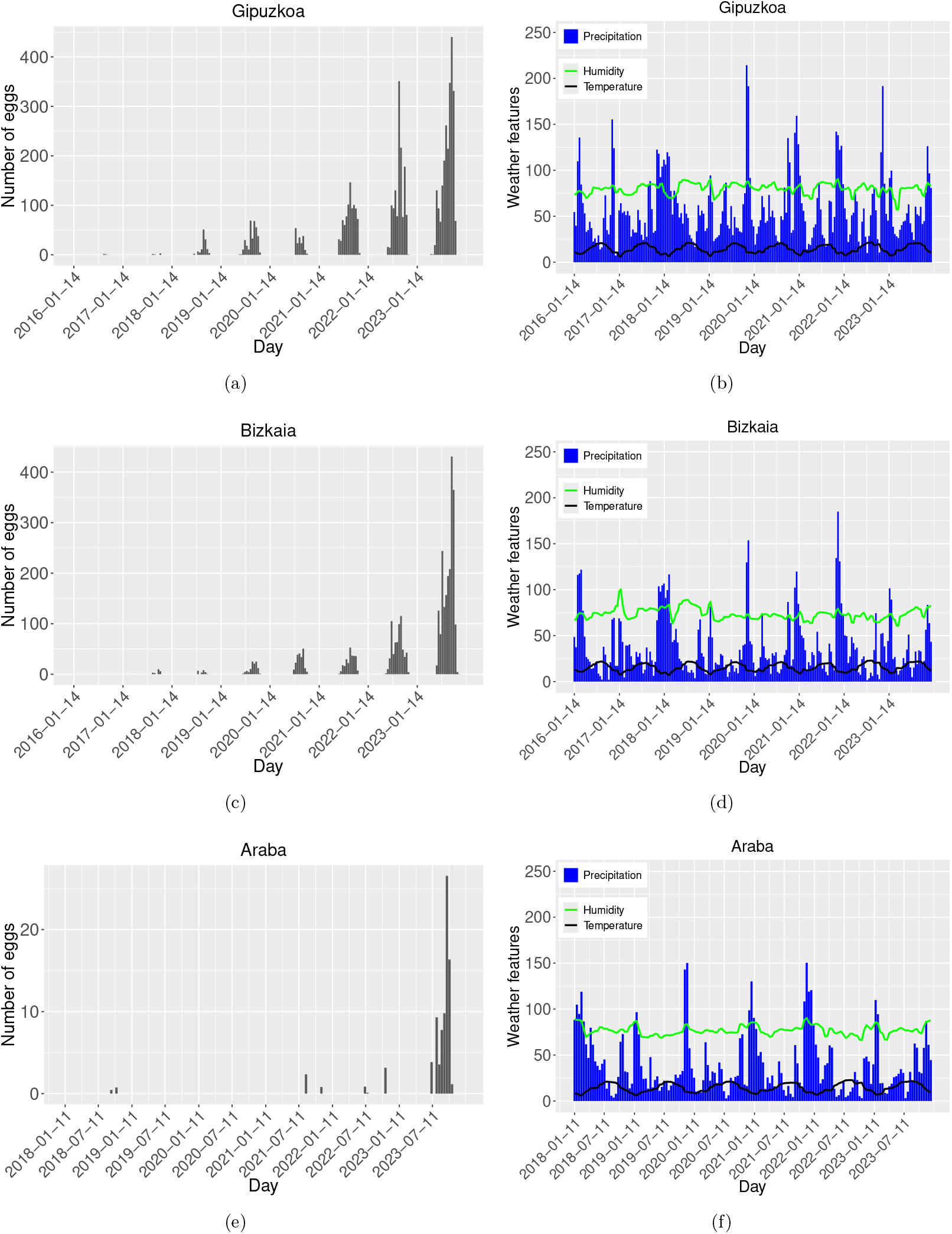
Number of mosquitoes eggs collected in (a), (c), (e). And average temperature (°*C*), relative air humidity (%), and cumulative precipitation (*mm*) in (b), (d), (f). Data gathered biweekly for Gipuzkoa, Bizkaia, and Araba, respectively.

Mosquito eggs are typically found during the summer months, from June to October, when the combination of higher temperatures and favorable humidity conditions promotes their activity and reproduction. As shown in Figure 2(a), the egg count in the entire Gipuzkoa province has significantly increased over the last years of collected data, although this trend may vary between municipalities. For example, in the city of Irun (see Supplementary Material B), the second most populated city in Gipuzkoa, located on the border with France, where variability is present without a clear increasing trend.

In Gipuzkoa, temperature exhibited a clear seasonal annual pattern, while accumulated rainfall showed no apparent trend. Humidity, however, decreased during the winter and followed a quasiperiodic structure (see Figure 2(b)). The winter of 2019, right after the expected period of higher egg presence, was exceptionally rainy compared to other winters in the province. Combined with low humidity (below 75%), this may have contributed to the lower egg counts observed in the following summer season (2020). In contrast, the dry summer of 2022, accompanied by higher humidity levels (above 75%), may explain the increased egg counts observed that year.

In Bizkaia, the time series of egg counts has displayed a consistent upward trend over the years, with positive egg traps first recorded in 2017 (see Figure 2(c)). Notably, the average mosquito egg count in Bizkaia during 2023 serves as a good proxy for the province-wide average, as shown by the time series for Bilbao, the capital of Bizkaia (see Figure 13(c) in in the Supplementary Material B).

The temperature in Bizkaia followed a clear seasonal pattern, while accumulated rainfall showed no apparent trend, with significant cumulative precipitation occurring later in 2021. In contrast to Gipuzkoa, however, humidity in Bizkaia exhibited periodic increases approximately every two years, with higher levels typically observed during winter months (see Figure 2(d)).

Moreover, average precipitation in Bizkaia was slightly lower than in Gipuzkoa. Temperature fluctuations in Bizkaia were more pronounced, as indicated by the steeper slope of its temperature curve compared to Gipuzkoa, potentially explaining the lower average egg counts in the region. Additionally, the dry summer of 2021, followed by a rainy winter, may have contributed to the consistent egg count trend observed.

Furthermore, although ovitraps have been distributed and data collected in the province of Araba since 2013, positive egg traps were not recorded until 2018, with no positive ovitraps observed in 2019 or 2020 (see Figure 2(e)). In Laudio, the second most populated municipality in Araba, positive ovitraps were only recorded in 2021 (see Figure 13(e) in Supplementary Material B).

The average temperature in Araba exhibits annual seasonality, while precipitation lacks a clear trend, though cumulative rainfall is typically higher during winter. On the other hand, humidity also tends to increase alongside precipitation (see Figure 2(f)). The lower average temperature in this province may contribute to the reduced presence of mosquito eggs.

Given the dispersed nature of data in Araba, with many zero values in egg counts (Figure 2(e)), there is insufficient information to develop a reliable training dataset for model fitting. Therefore, this province is excluded from further analysis. Smaller spatial units, such as individual municipalities, are similarly excluded, with the focus of this study being the two Basque Country provinces, Gipuzkoa and Bizkaia. Nonetheless, descriptive statistics and detailed analyses at the municipal level for Irun and Bilbao, which have adequate data, are provided in the Supplementary Material B.

### 2.2 Methodological approach

#### 2.2.1 Data processing

After gathering data, pre-processing is a crucial initial step before model training, forecasting, and evaluation. In this study, data pre-processing included the following steps. First, we ensured a consistent interval for both the independent and dependent variables, selecting a biweekly interval for the entomological data based on the average 14-day period in which egg counts were collected.

Next, we addressed missing values through imputation, filling gaps with zero values. This choice is scientifically justified within the context of this dataset, as institutional data indicated that, for months without data collection, ovitrap counts would have likely been zero [9]. This assumption was based on data from four sentinel points (two in Gipuzkoa and two in Bizkaia) monitored over a year to determine the start and end of *Aedes* mosquito activity in regions with recorded presence in the previous year.

Additionally, we included only the 20 highest egg counts at the provincial level to account for variations in the number of monitored ovitraps over time, helping to reduce dataset skewness. Outliers were then removed using a central moving average as a smoothing method, commonly applied to mitigate white noise, random fluctuations, and extreme values [39].

For the meteorological data, no imputation was required as daily weather data was available for the entire study period. In this case, outliers were retained as they could signal significant events associated with the presence or absence of mosquito eggs. Basic exploratory analysis was then conducted using descriptive statistics and correlation tests, incorporating both the original and lagged versions of the meteorological data.

Finally, we split the data into training and testing sets, with the training data comprising 85.71% and 83.33% for Gipuzkoa and Bizkaia, respectively. The remaining 26 data points (one year of biweekly data) were allocated for testing.

#### 2.2.2 Models

In this study, we applied different models including and excluding the lagged version of eggs count as a proxy and the lagged version of the independent environmental variables. To appropriately handle the discrete and non-negative nature of counts, we restrict our choices and applications of the models presented here. For instance, the statistical methods such as the Poisson Regression and Negative Binomial Regression are foundational models for count data [36]. However, while the first one assume that the time series follows a Poisson distribution, the second one can be useful when time series presents more variability and over-dispersion (i.e., the variance is greater than the mean) (as it is the case). Both models are an extension of the Generalized Linear model (GLM) with a log link function.

The GLM is a flexible extension of ordinary linear regression that accommodates response variables with error distributions other than the normal distribution. This model often outperforms others when applied directly to the original data, compared to the transformed data such as using logarithmic scale [2, 30]. As such, we initially avoided any normalization or transformation of the data. When we applied the GLM to this dataset, it performed better on the smoothed data (using a three-point central moving average) than on the original, unprocessed data. And, a GLM with the canonical link function was used, assuming a Gaussian distribution for the response variable. In other words, the response variable follows a Gaussian exponential family distribution. This allows for more flexibility in modeling, as it does not impose the strict relationship between mean and variance required by models such as when using the Poisson distribution [23].

The GLM with a Gaussian family assumes a linear relationship between the predictors and the response variable **Y**, using the identity link function. That is, the conditional mean ***μ*** is a linear combination of unknown parameters ***β*** via the link function *g*:

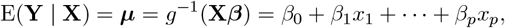

where E(**Y** | **X**) is the expected value of **Y** conditional to **X**, and *g* is the link function, which in this case, is the identity function [23]. The model predicts the mean of the response variable based on the input variables, and estimates the coefficients ***β*** by maximizing the likelihood function, assuming that the residuals are normally distributed.

Moreover, not only the dataset is over-dispersed but the response variables contains a lot of zeros, due to two main reasons: one the absence of positive eggs count outside during winter season (the so-called true negative) and the absence of more samples in more localities (the so-called false negative). In this case, zero-inflated models can handle excess zeros effectively. Zero-Inflated Negative Binomial (ZINB) can be effective in this case assuming that the data come from a mixture of two processes: one generating zeros and another generated by a negative binomial distribution [37]. Other models that can handle over-dispersion and the zero counts is the Generalized Additive Models for Location, Scale, and Shape (GAMLSS) which is flexible in modeling different distributions, not just the mean but also the variance [45]. However, after applying these models to the dataset, we observed that they were prone to over-fitting, indicating that the model might learned the noise in the training data rather than generalizing the unseen data.

On the other hand, our predictors are temporal series mostly exhibiting seasonal trends. Time series models like Seasonal Autoregressive Integrated Moving Average (SARIMA) are commonly used for forecasting since it is suitable for temporal count data and can handle seasonality. SARIMA has been extended to the Seasonal Autoregressive Integrated Moving Average with Exogenous variables (SARIMAX) which can include exogenous variables giving more accurate outcomes. SARIMAX combines differencing, autoregression, moving averages, and seasonal components, incorporating as well exogenous predictors [24]. Unlike models such as the GLM, this model assumes that the response variable depends on its past values *Y*_*t*_. Also, of its past forecast errors *ϵ*_*t*_, and external predictors, capturing temporal effects. This relation reads:

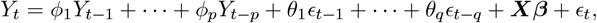

where *Y*_*t*_ is the response variable at time *t, ϕ*_*i*_ are the the autoregressive (AR) parameters, *θ*_*i*_ the moving average (MA) parameters, ***X*** is the predictors (exogenous variables), and ***β*** is the vector of coefficients.

The model depend, as well, on the order of the AR terms *p*, representing the number of lagged values of the series used in the model; the degree of differencing *d*, which removes trends and makes the series stationary; the order of the MA terms *q*, representing the number of lagged forecast errors. And, on the seasonal component, *P, D*, and *Q* that are the seasonal terms for the parameters *p, d*, and *q*, respectively, and on is the length of the seasonal cycle *s* [24]. We implemented SARIMAX in the R compute language by using the `auto.arima()` function that automatically selects the best seasonal and non-seasonal parameters *p, d, q, P, D, Q*, and *s* based on the data.

On the other side, machine learning techniques such as Random Forest (RF) [6], Conditional Inference Trees (CTree), and Artificial Neural Networks (ANNs) can be also used for forecasting count data. However, in this study, ANNs is the least performing machine learning model, a finding corroborated by previous research [37] which do not advises using ANNs for count data with over-dispersion.

RF builds decision trees using bootstrap samples and random feature subsets, and combines the predictions from all trees. Each tree is developed using a subset of features, as chosen by the `mtry` parameter [6]. The `mtry` parameter determines the number of predictor variables considered at each split, playing a crucial role in controlling over-fitting. The `ntree` parameter refers to the number of trees to be generated in the RF. Increasing the number of trees generally enhances model stability and robustness, although beyond a certain threshold number of trees, the additional trees yield to insignificant improvements in the terms of model performance. The advantage of RF lies in its ability to handle complex data and it is designed to mitigate over-fitting [14, 37].

The final RF predictions for the conditional mean of ***Y***, given the predictor ***X***, is based on the average or weighted average of all the individual trees’ predictions. Thus, the RF model can be expressed as:

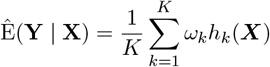

where *h*_*k*_(***X***) is the prediction of the k-th tree, and *K* is the total number of trees [6]. Each tree is built using a bootstrap sample of the original data and selects features at random from the `mtry` subset.

On the other hand, the CTree is a non-linear method to model the relationships between predictor variables and a response variable. The CTree algorithm recursively partition the dataset based on the values of the predictors, using statistical tests to determine the significance of potential splits. The splits are chosen by testing the association between each predictor and the response, and the predictor with the strongest association (lowest p-value) is selected for each split.

The conditional distribution of ***Y***, given the predictor ***X***, is estimated as:

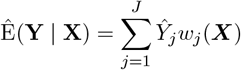

where *Ŷ*_*j*_ is the predicted value for the j-th terminal node, *w*_*j*_(***X***) is the weight indicating whether observation *j* falls into the same terminal node as ***X*** [42].

While RF and CTree both rely on decision tree methodologies, they differ in their approaches. RF employs random feature selection to create an ensemble of trees, which enhances generalization but sacrifices interpretability. Conversely, CTree focuses on unbiased variable selection, offering better interpretability. RF generally offers better predictive performance on large and complex datasets, while the structural differences in the partitions can highlight the unique advantage of CTree.

We implement the GLM, SARIMAX, RF and CTree models (and other models discussed in this section) in the R computing language (R version 3.6.3) using the packages `MASS, forecast, randomForest` and `party`, respectively. Nevertheless, only the four models discussed earlier will be presented in this study because, as previously mentioned, some models exhibit over-fitting, others demonstrate under-fitting (as is the case with the ANNs model), and some fail to capture any significant features of the data.

#### 2.2.3 Stationary analysis

We applied the augmented Dickey-Fuller (ADF) test, a commonly used method for testing the presence of a unit root in time series data, to assess whether the time series is non-stationary. Non-stationarity in a time series often presents means, variances, and covariances that change over time, making the series unpredictable and challenging to model or forecast. Although some models, such as SARIMAX, can handle non-stationarity, stationary time series often yield more reliable results. The null hypothesis of the ADF test states that the series contains a unit root, indicating non-stationarity, while the alternative hypothesis suggests that the series is stationary. To test the null hypothesis, we computed the *p*-value. A *p*-value less than 0.05 leads us to reject the null hypothesis, implying stationarity.

We conducted the ADF test using the `tseries` package in R. For both datasets, Gipuzkoa and Bizkaia, the ADF test on the predictor variable yielded a *p*-value of approximately 0.01 < 0.05, indicating that the datasets are stationary.

#### 2.2.4 Evaluation metrics

To compare the performance of statistical and machine learning models, three widely used evaluation metrics were employed: the Mean Absolute Error (MAE), the Root Mean Squared Error (RMSE), and the R-squared (*R*^2^) score.

The MAE is calculated as:

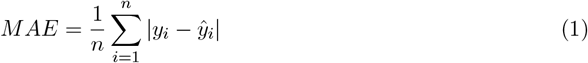

where *y*_*i*_ and *ŷ*_*i*_ represent the observed and predicted values, respectively, and | *·* | denotes the absolute value [37]. MAE measures the average magnitude of the errors in a set of predictions, without considering their direction.

The RMSE is given by:

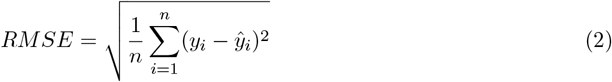

where *y*_*i*_ and *ŷ*_*i*_ are the observed and predicted values, respectively. RMSE gives a higher weight to large errors compared to MAE and is sensitive to outliers.

The *R*^2^ score, also known as the coefficient of determination, is calculated as:

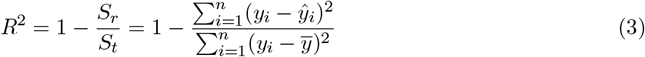

where, *S*_*r*_ is the Residual Sum of Squares, representing the sum of squared differences between the observed values (*y*_*i*_) and the predicted values (*ŷ*_*i*_); and *S*_*t*_ is the Total Sum of Squares, calculated as the sum of squared differences between the observed values (*y*_*i*_) and their mean (*y*). An *R*^2^ score of 1 indicates that the model explains all the variability of the response variable, while a score of 0 indicates no explanatory power.

The selection of the best model is based on achieving the lowest MAE or RMSE values, or an *R*^2^ score closest to 1. In this study, the MAE is chosen as the primary evaluation metric due to its suitability for machine learning models [37].

## 3 Results

### 3.1 Exploratory statistical analysis

Basic exploratory statistical analysis was performed, starting with descriptive statistics for both the response and predictor variables (after pre-processing and smoothing). All variables in the dataset were found to be skewed and over-dispersed. The null hypothesis of normal distribution was rejected based on the results of the Kolmogorov-Smirnov test and the Shapiro-Wilk test, both of which yielded *p*-values < 0.05, indicating significant deviation from normality for all variables.

For the Gipuzkoa dataset:

- Eggs count had a mean of 25 and a median of 0.
- Temperature (in °C) had a mean of 14.6 and a median of 14.2.
- Relative air humidity (in %) had a mean of 79.9 and a median of 80.8.
- Precipitation (in mm) had a mean of 57.6 and a median of 49.4.

For the Bizkaia dataset:

- Eggs count had a mean of 16 and a median of 0.
- Temperature (in °C) had a mean of 15.5 and a median of 14.8.
- Relative air humidity (in %) had a mean of 73.6 and a median of 72.8.
- Precipitation (in mm) had a mean of 37.4 and a median of 27.1.

Additional details are provided in the Supplementary Material (see Figure 10), as well for the province of Araba.

The relationship between meteorological variables and the number of mosquito eggs was explored using scatter plots (see Figures 3(a)-(c) for Gipuzkoa and 3(d)-(f) for Bizkaia). No linear relationship was confirmed, as indicated by Pearson’s correlation index. Nevertheless, it is well-known that the combination of high temperatures (22 °C to 27 °C) and high humidity increases oviposition rates (egg-laying) in adult female mosquitoes [20, 25]. This association is reflected in Figures 3(a) and (b) for Gipuzkoa, and Figures 3(d) and (e) for Bizkaia.

**Figure 3:**
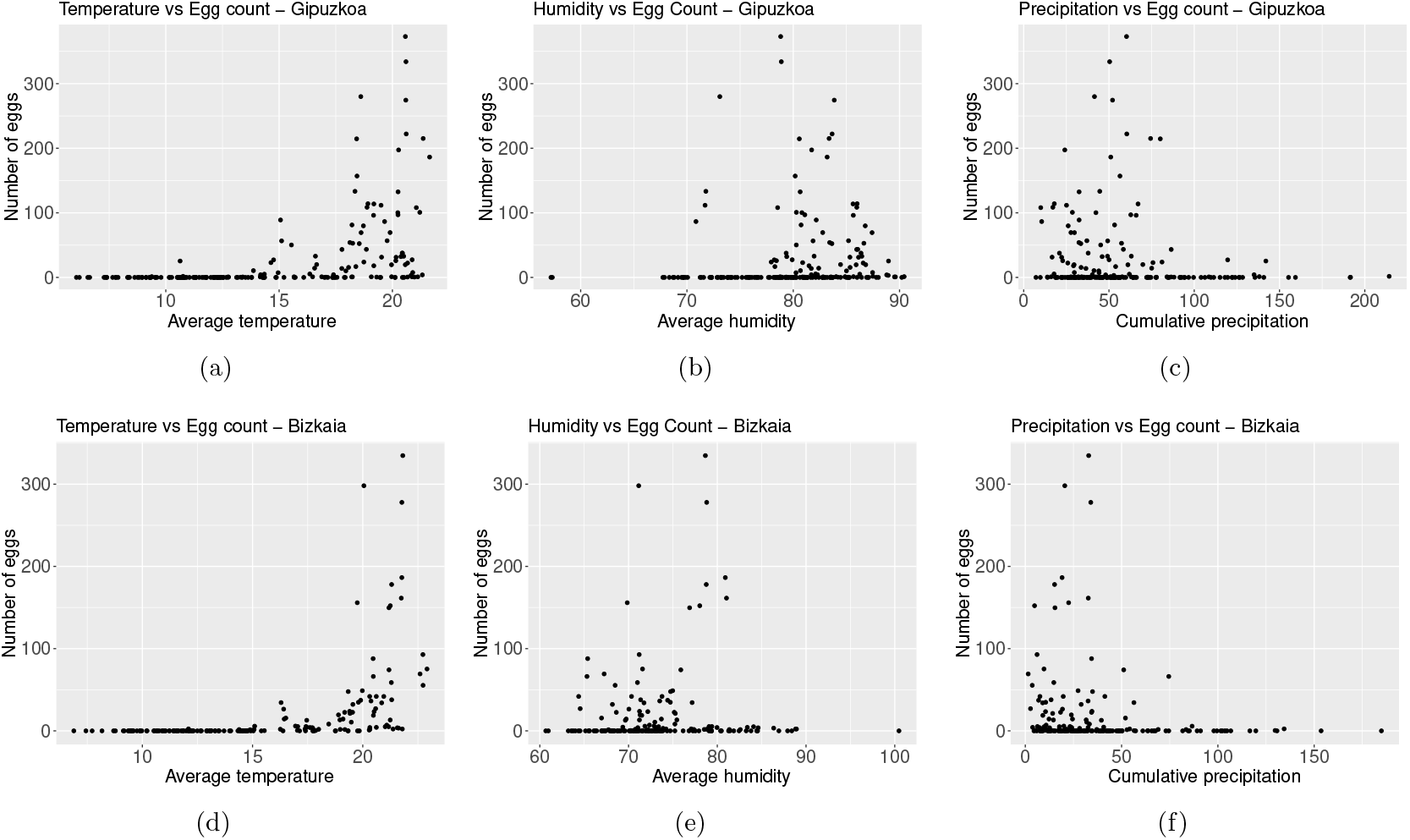
Average temperature (in °C) versus the number of collected mosquito eggs, in (a), (d). Average relative air humidity (in %) versus the number of collected mosquito eggs, in (b), (e). Accumulated precipitation (in mm) versus the number of collected mosquito eggs, (c), (f). Data gather biweekly, in Gipuzkoa and Bizkaia, respectively.

As described in Section 2.2.1, the time series data were smoothed using a central moving average to reduce short-term fluctuations and noise. This preprocessing step helped mitigate spurious short-term correlations and revealed underlying long-term relationships between variables, thereby increasing correlation indexes.

On the other hand, Spearman’s correlation analysis confirmed a strong monotonic relationship between the number of eggs and temperature, with a correlation index ≥ 0.72 (see Figures 4(a) for Gipuzkoa and 4(b) for Bizkaia). Although no significant correlations were found between egg counts and the other climate variables, the direction and strength of these relationships are displayed in Figures 4(a) and 4(b). Specifically, As humidity increases, the number of eggs increases, showing an intermediate correlation. In contrast, as accumulated precipitation increases, the number of eggs decreases, albeit with very low or negligible correlation.

**Figure 4:**
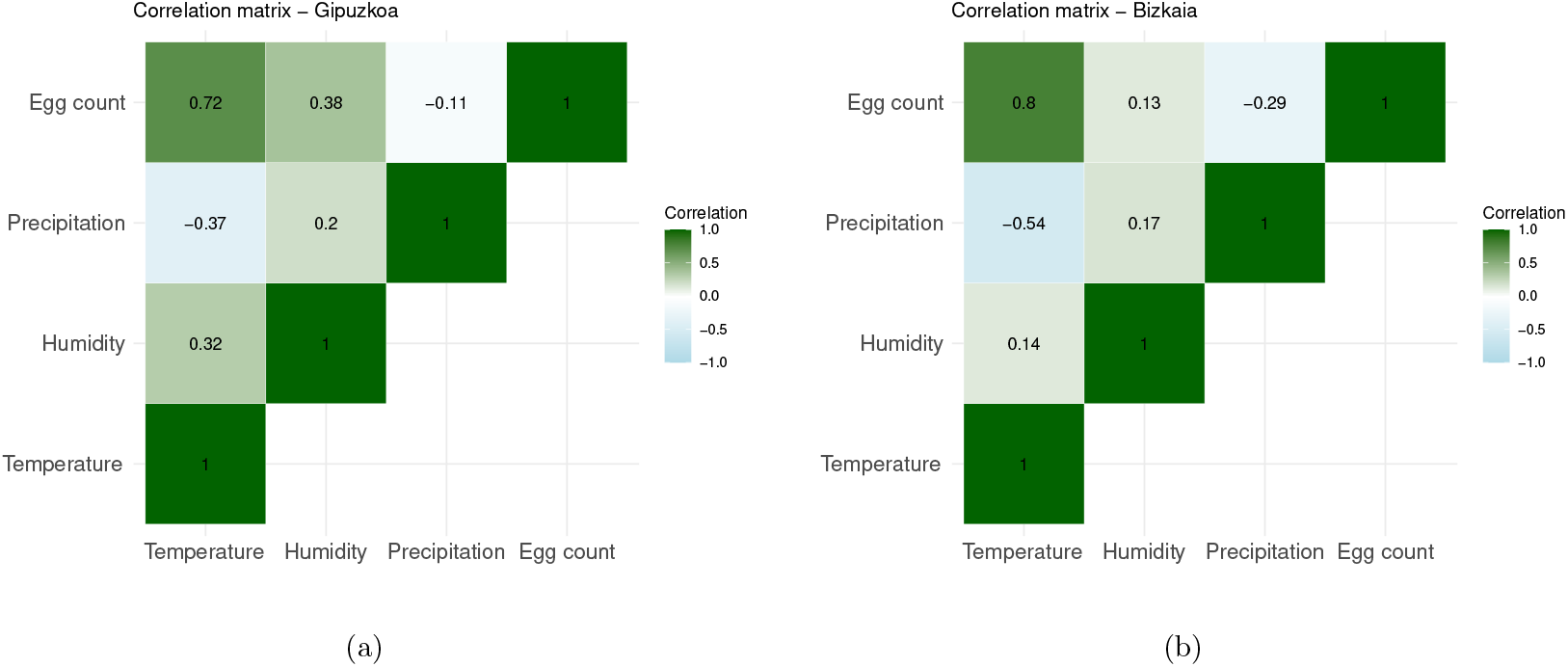
Spearman correlation matrix between weather features and the number of mosquito eggs. The matrix shows a high correlation between the number of eggs and temperature (index = 0.72 for Gipuzkoa and index = 0.8 for Bizkaia), but no significant correlation with the other features.

In addition, we have created lagged time series for all the meteorological variables (see Figure 5(a)-(c) and 5(d)-(f)), and we evaluate the monotonic correlation using Spearman correlation, highlighting the time lag at which the highest index value occurs (see Figure 11(a) and 12(a) in the Supplementary Material A). At the time lag at which the highest correlation value occurs, the lagged time series will be used as predictor variables (see Figure 11(b) and 12(b) for Gipuzkoa and Bizkaia, respectively, in the Supplementary Material A).

**Figure 5:**
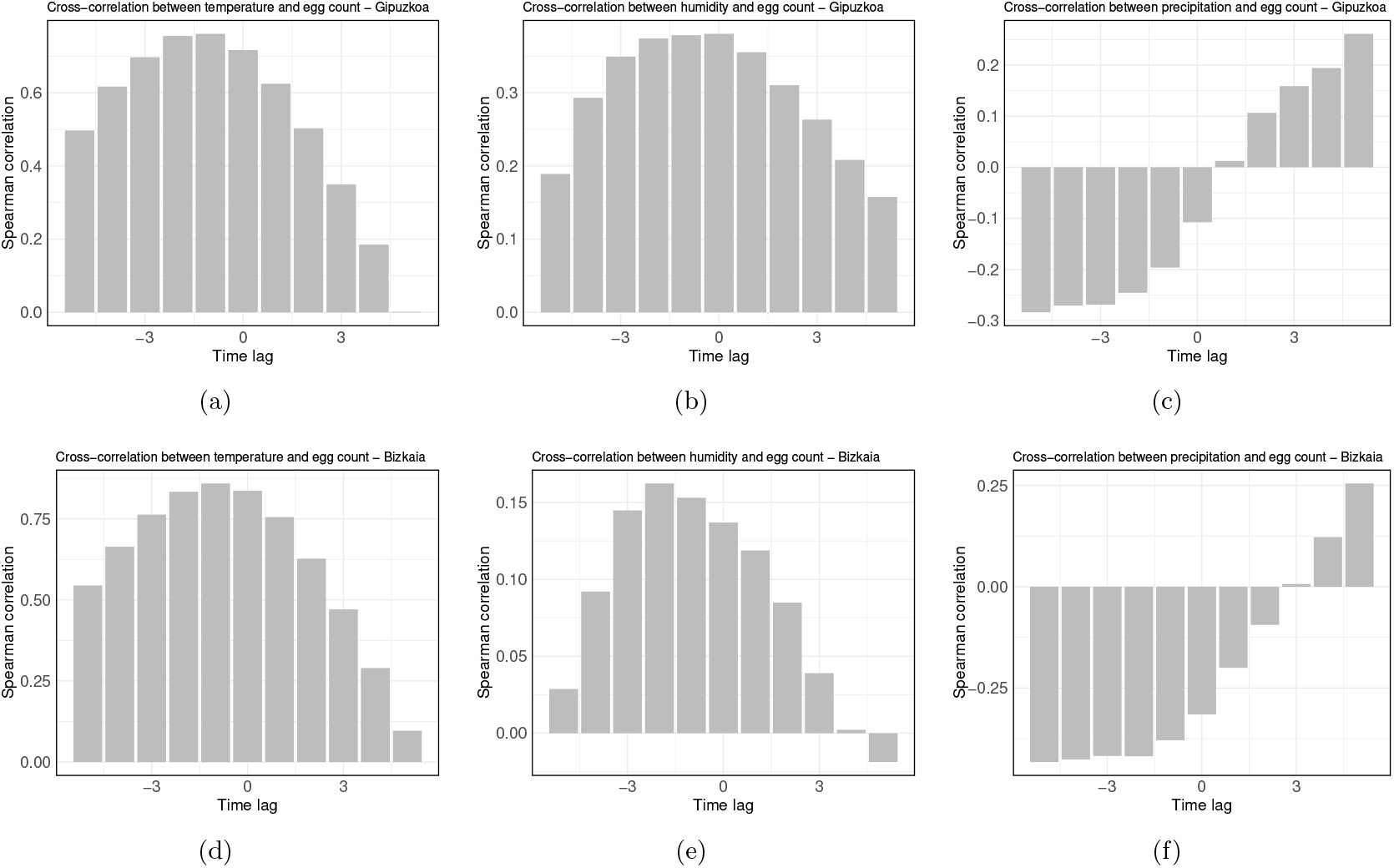
Spearman correlation between the lagged time series of weather features and the number of mosquito eggs, with a time lag of 1 unit (2 weeks). For temperature, the maximum correlation occurs at a lag of −1 unit, in (a), (c). For humidity, the maximum correlation occurs at a lag of 0 units and −2 units, for Gipuzkoa and Bizkaia, in (b), (e) respectively. For precipitation, the maximum correlation occurs at a lag of −5 units, in (c), (f).

For instance, Figure 5(a) shows that the highest correlation value between egg counts and temperature series, in Gipuzkoa, occurs at lag -1. This could imply that the egg production series is most strongly correlated with the temperature series 2 weeks (1 period) earlier. Therefore, changes in temperature might have a leading effect on egg production, where temperature changes influence egg production with a delay of 1 period (2 weeks). For humidity (Figure 5(b)), the highest correlation occurs at 0 units (0 weeks) with a (low) positive correlation, while for precipitation, (Figure 5(c)), the highest correlation occurs at lag -5 units (10 weeks) with a negative (low) correlation.

For Bizkaia, Figure 5(d) shows that the highest correlation value between egg counts and temperature series occurs at lag -1. This could imply that the egg production series is most strongly correlated with the temperature series 2 weeks (1 period) earlier. Therefore, changes in temperature might have a leading effect on egg production, where temperature changes influence egg production with a delay of 1 period (2 weeks). For humidity, (Figure 5(e)), the highest correlation occurs at lag -2 units (4 weeks) with a positive correlation, while for precipitation, (Figure 5(f)), the highest correlation occurs at lag -5 units (10 weeks) with a negative correlation.

High correlation is shown between egg counts and temperature at lag -1, with an index value of 0.76 and 0.83 for Gipuzkoa and Bizkaia, respectively (see Figures 11(b) and 12(b) in the Supplementary Material A). While intermediate to low correlation appears to be positive and correlated between humidity and egg counts, Figures 3(b) and 3(e) show that the highest egg count occurs when humidity percentages are between 70% and 80%.

Moreover, a low negative correlation between precipitation and egg counts was found (see Figures 5(c) and 5(f)). Although the strength of the correlation is considered low, the opposite direction in the correlation for precipitation approximately 10 weeks prior to egg collection (almost 3 months earlier) can be explained by the fact that periods of high precipitation temporarily reduce the number of females actively searching for a host and, therefore, laying eggs [18]. On the other hand, drier periods occurring 10 weeks before the collection, increase the egg counts. This can be attributed to the fact that mosquito eggs are extremely resistant. They can remain viable in a dry state within a container for 300 to 400 days without direct water contact, allowing them to stay in ovitraps for extended periods without hatching [31].

### 3.2 Fitting and error analysis

Prior to model fitting, the dataset was divided into training and testing sets. For Gipuzkoa, the training dataset includes data from 2017 to 2022 (85.71%), while the test dataset consists of data points from the year 2023 (see Figure 6). In contrast, for Bizkaia, the training dataset covers the period from 2018 to 2022 (83.33%), with 2023 as the test dataset (see Figure 7). The choice of yearly data is related to the frequency of data availability, while the starting point corresponds to the need for cleaning the missing values (NA not available) due to the lagged versions of variables.

We train several models on the training dataset, considering, lagged version of the independent variables, as well as including and excluding lagged version of the eggs count variable. The majority of models performed better including the proxy lagged version. Here we include only models with the best performances. Which are: the Random Forest (RF) model, the Generalized Linear Model (GLM) with Gaussian distribution (here abbreviated by GLMG), the Seasonal Autoregressive Integrated Moving Average with Exogenous variables (SARIMAX) model and, the Conditional Inference Trees (CTree) (here abbreviated by CT).

**Figure 6:**
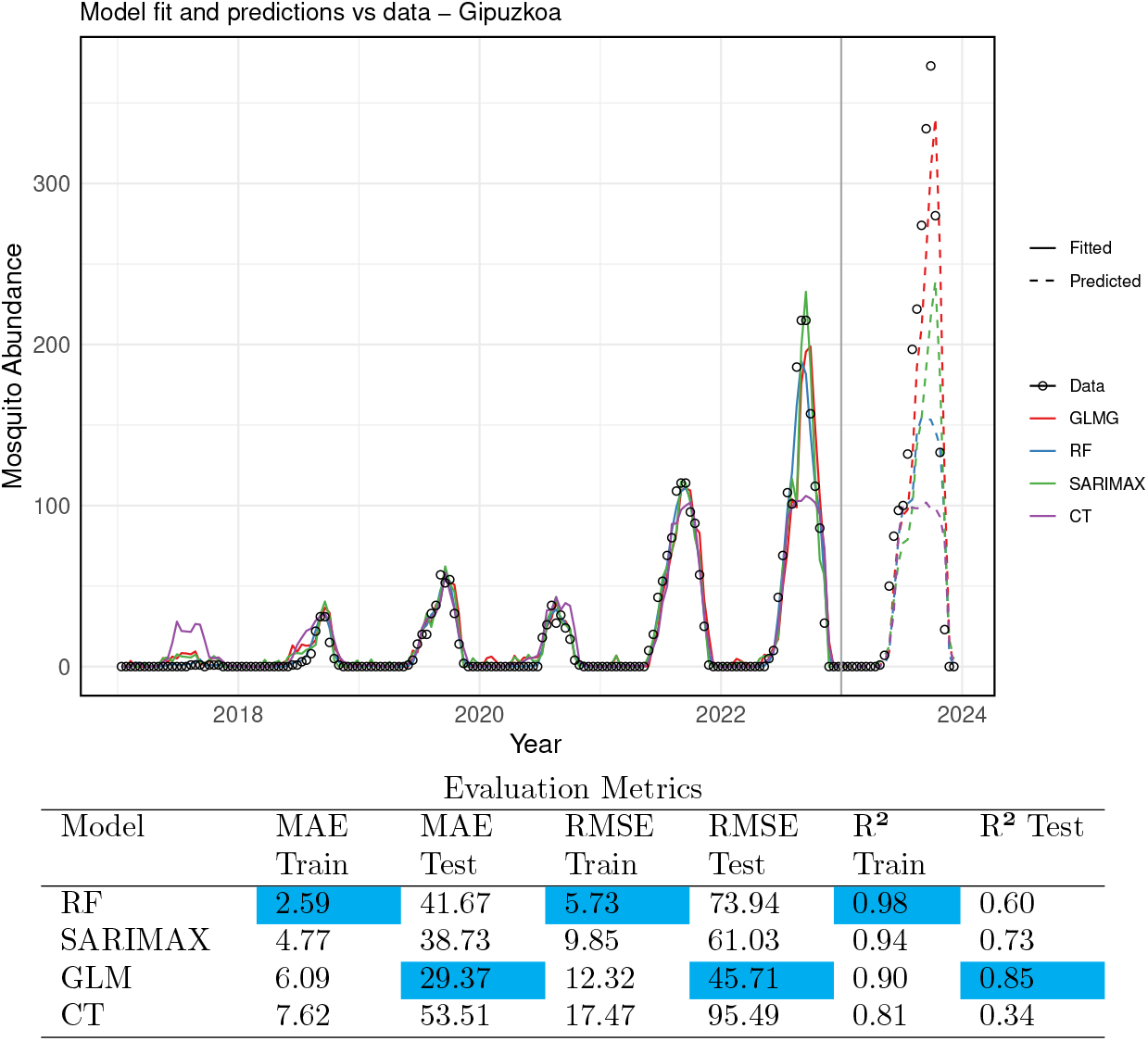
Comparison of actual data with the fitted and test values for Gipuzkoa. The actual data is represented by open black circles, while the fitted values are shown as solid lines and the test values as dashed lines. The models are represented as follows: in blue, the Random Forest (RF) model (`ntree` = 600, `mtry`= 5); in red, the Generalized Linear Model (GLMG); in green, the SARIMAX model; and in purple, the Conditional Inference Trees (CT) model (`ntree` = 500, `mtry` = 3). The vertical gray line separates the training dataset (2017–2022) from the test dataset (2023). The table shows the error metrics for each chosen model, on the training and test datasets for Gipuzkoa.

**Figure 7:**
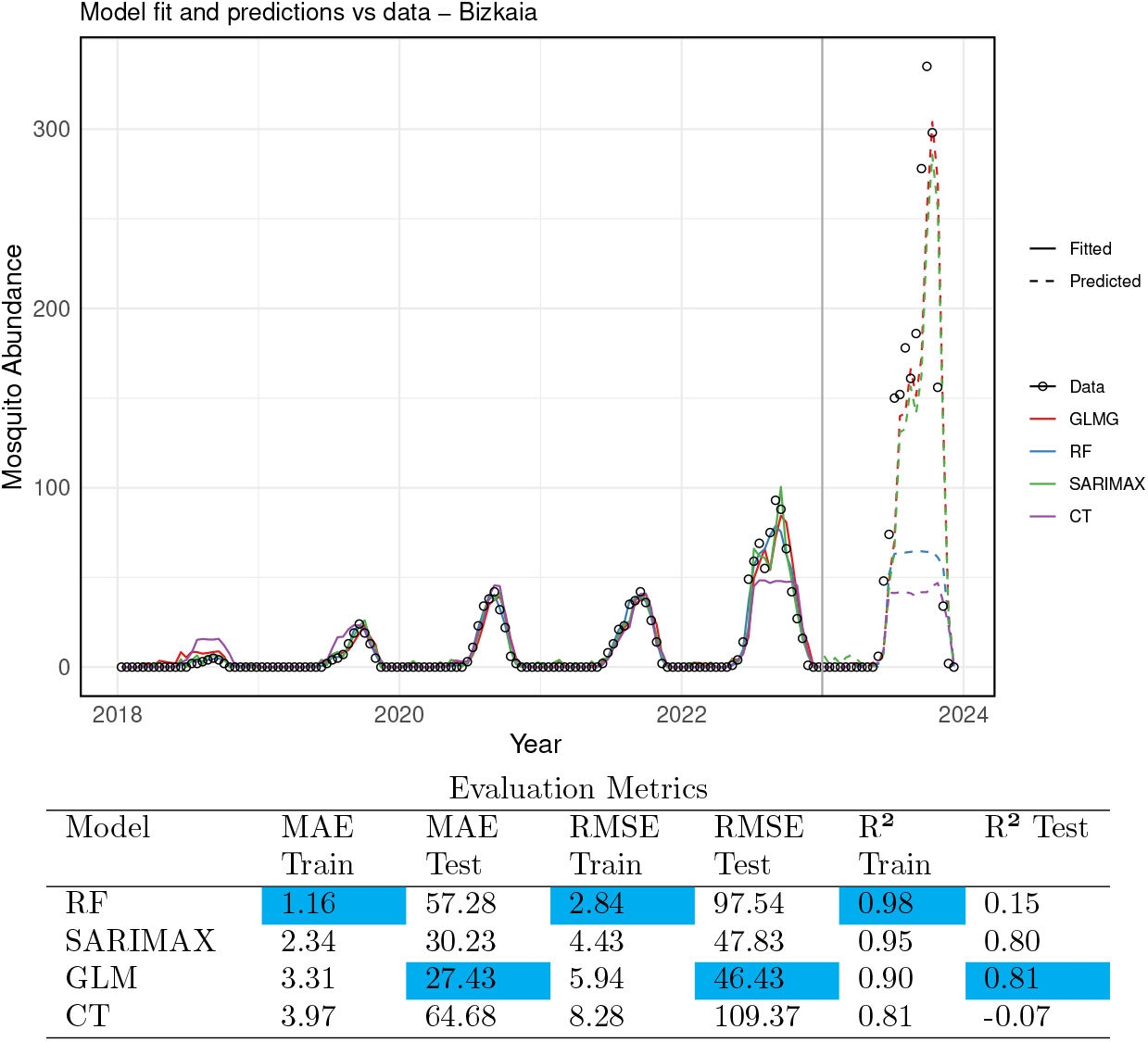
Actual data versus the fitted and test values of the models for Bizkaia. The actual data is represented by open black circles, while the fitted values of each model are shown with solid lines, and the test values with dashed lines. In blue, the RF model (`ntree` = 600, `mtry` = 5); in red, the GLMG; in green, the SARIMAX model; and in purple, the CT model (`ntree` = 500, `mtry` = 3). The vertical gray line delineates the training dataset (2018–2022) from the test dataset (2023). The table shows the error metrics for each chosen model, on the training and test datasets for Bizkaia.

We implement the GLMG, SARIMAX, RF and CT models in the R computing language (R version 3.6.3) using the `glm(), auto.arima(), randomForest() and cforest()` function, respectively. We employed and compare the models on the training dataset and on the testing dataset. Later, we evaluate each models performance in the datasets using MAE, RMSE and *R*^2^ metrics.

Based on the evaluation metrics the best performance on the training dataset for Gipuzkua is the RF model. The model could explain 98% of the variance in the data, according to the *R*^2^ evaluation While in the test dataset 60%. For the test dataset GLMG was the model that performed better, explaining 85% of the variance in the dataset, followed by the SARIMAX model (see Figure 6).

Although the RF model performed best during training, its predictions ranked in third place, which might suggests an over-fitting. On the other hand, even though GLMG did not top the training performance, it gave better predictions, making it a more reliable model overall. This suggests that the simplicity of GLMG helped it generalize better to the unseen data, while RF may have captured the noise from the training set, which could reduce the predictive accuracy.

In the case of Bizkaia, the RF model performed best on the training dataset, explaining 98% of the variance, as indicated by the *R*^2^ value. However, on the test dataset, it explained only 15% of the variance. For the test dataset, the GLMG model performed the best, explaining 81% of the variance, closely followed by the SARIMAX model with 80% (see Figure 7).

Among the four models evaluated, the CT model performed the worst, based on all error metrics for both the training and test datasets. Additionally, the CT model was unable to explain the test dataset for Bizkaia.

The poor performance of the models on the Bizkaia test set, as shown in the time series, can be attributed to differences in the characteristics of the training and test data. Notably, the mean value of the training data is significantly lower than that of the test data, with egg counts in 2023 being unusually high. This discrepancy between the training and test datasets likely contributed to the models’ suboptimal performance for Bizkaia.

After training, testing, and evaluating each model, we used the models with the best performance to predict future *Aedes* invasive mosquito abundance. For this, we included 2023 data points in the training dataset and, using the historical time series data along with lagged versions of the variables, we forecast values based on the last observations.

Figure 8 shows the fitted values and predictions for mosquito abundance in Gipuzkoa for 2024, while Figure 9 presents the same for Bizkaia. Only the three models with the best performance are displayed. It is noteworthy and expected that extending the training dataset length improved the performance of all models. This highlights the importance of maintaining entomological surveillance for more accurate future predictions.

**Figure 8:**
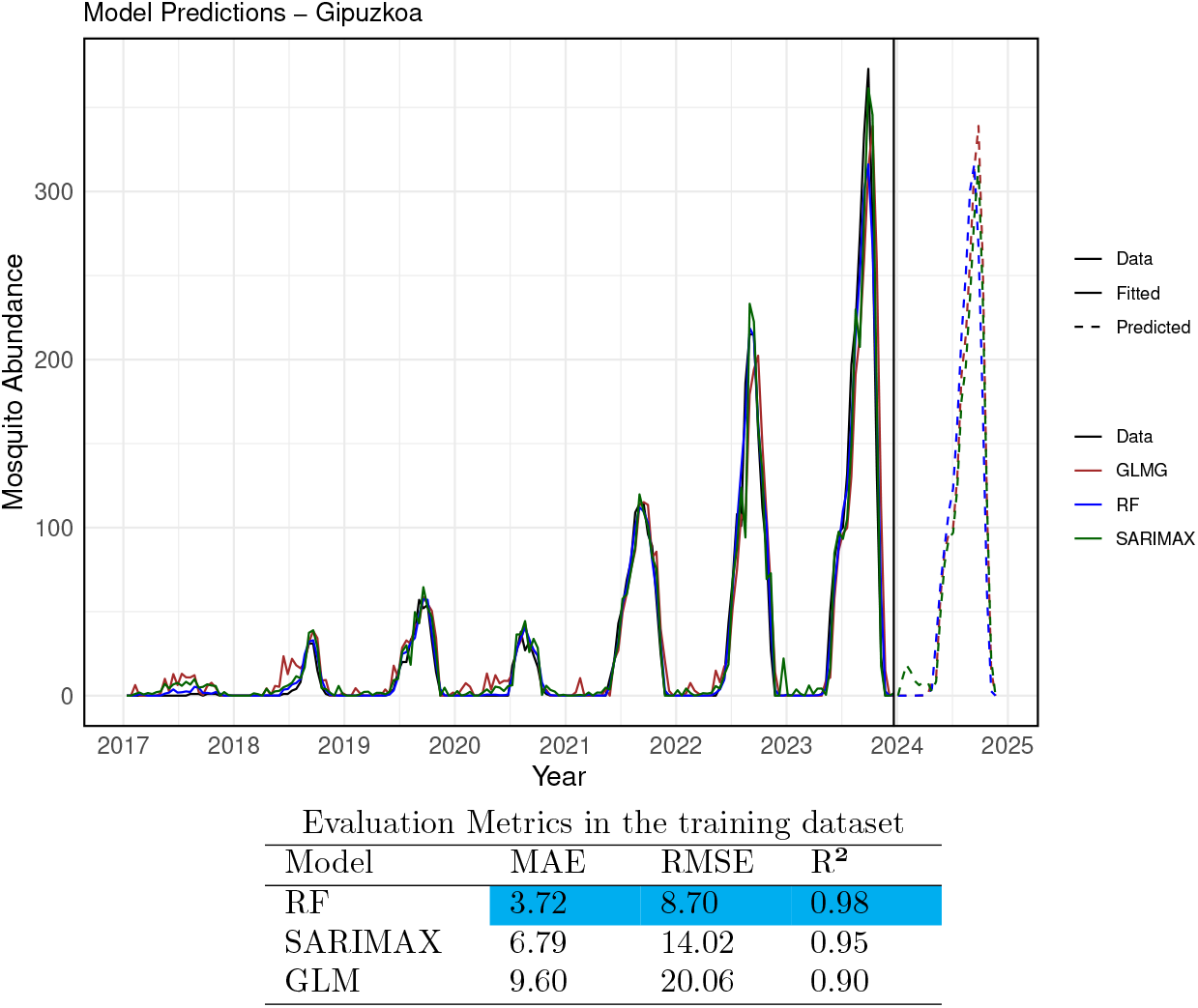
The actual data versus the fitted and predicted values for Gipuzkoa. The actual data is represented by a solid black line. The fitted values for each model are shown as solid colored lines, and the predicted values are displayed as dashed lines. In blue, the RF model (`ntree` = 600, `mtry` = 5); in brown, the GLMG model; and in green, the SARIMAX model. The vertical black line delineates the training dataset (from 2017 to 2023) from the forecasted period for the year 2024. The table shows Evaluation of error metrics in the training dataset (for Gipuzkoa) showing the best model performance.

**Figure 9:**
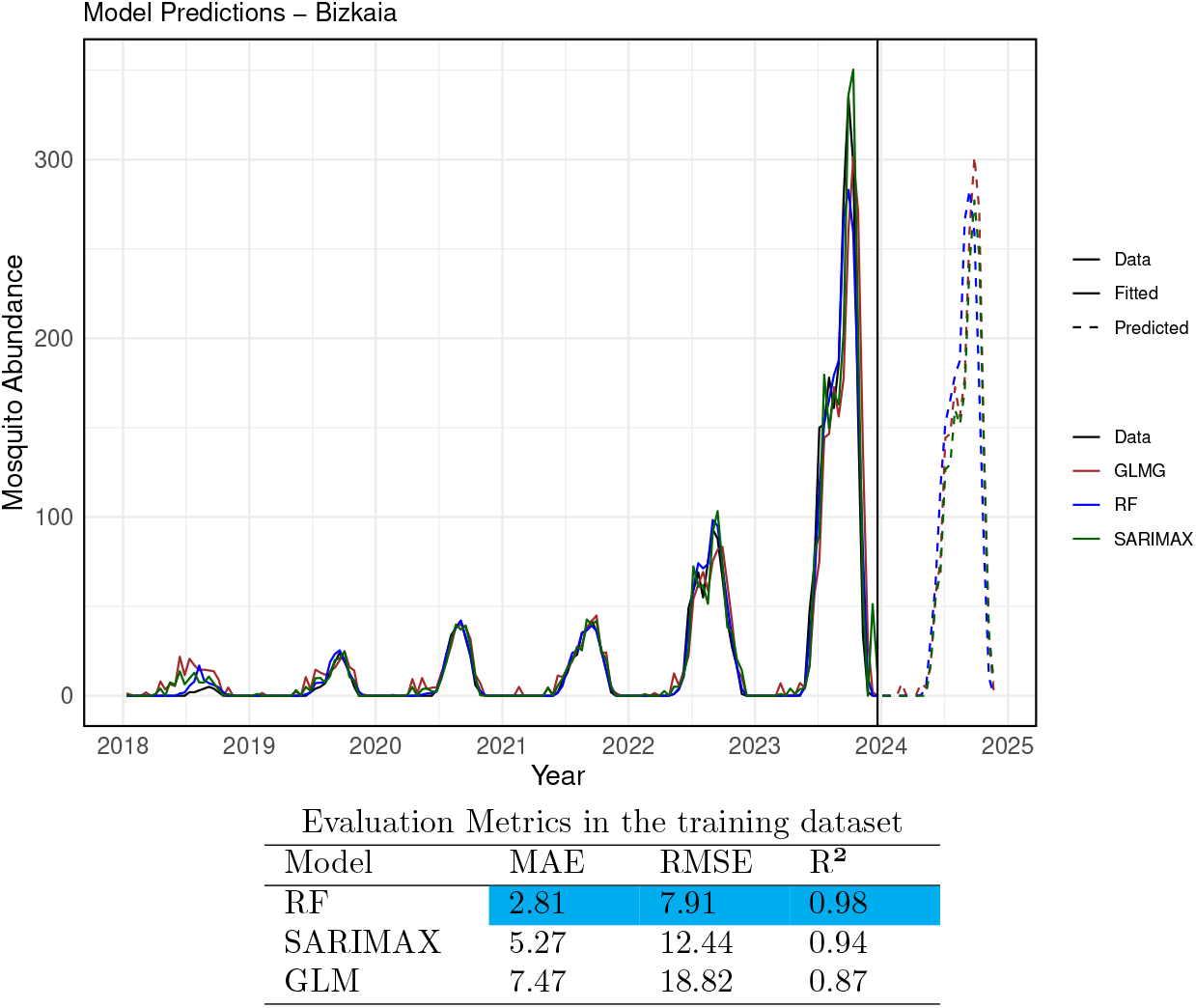
The actual data versus the fitted and predicted values of the model for Bizkaia. The actual data is represented by a solid black line, while the fitted values of each model are shown as solid colored lines, and the predicted values as dashed colored lines. In blue, the RF model (`ntree`= 600, `mtry` = 5); in brown, the GLMG model; and in green, the SARIMAX model. The vertical black line delineates the training dataset (from 2018 to 2023) from the forecasted period for the year 2024. The table shows the evaluation of error metrics in the training dataset (for Bizkaia) showing the best model performance.

The error analysis for the training dataset is shown in Figure 8 and Figure 9. The results indicate that, across different metrics, the RF model provided the best fit, explaining 98% of the variance in both Gipuzkoa and Bizkaia, making it suitable for forecasting. The SARIMAX model also performed well, explaining 95% and 94% of the variance in Gipuzkoa and Bizkaia, respectively.

At the municipal level (see more details in Supplementary Material B), the RF model performed best for Irun (in Gipuzkoa), while for Bilbao (in Bizkaia), the SARIMAX model provided the best fit, explaining 97% of the variance in the training dataset.

Furthermore, we estimate and expect that mosquito abundance in 2024 will be lower compared to the previous year, both at the provincial and municipal levels. This reduction may be due to various factors, such as changes in optimal environmental conditions and potential variations in weather patterns.

## 4 Discussions and conclusions

The Basque Country, an autonomous community in northern Spain, has experienced an increase in imported cases of mosquito-borne diseases, along with the establishment and expansion of *Aedes albopictus* and *Aedes japonicus* mosquitoes. This study uses egg count data retrieved from ovitraps monitored by the regional surveillance program conducted by the Department of Public Health of the Basque Government and the public agency NEIKER at various locations across the Basque provinces. We employ statistical models and machine learning techniques to model the relationship between the recorded mosquito ovitrap egg counts and climate factors such as temperature, humidity, and precipitation.

Before selecting the model, statistical analysis was conducted on the dataset to examine the influence of environmental factors on the predictor variables. We compared different models, including versions with and without lagged egg counts as a proxy. Importantly, incorporating lagged versions of all independent and dependent variables improved the performance of most models.

We found that forecasting mosquito abundance is particularly challenging in non-endemic areas, where no local mosquito-borne cases have been reported. While environmental factors are the primary drivers of mosquito abundance and distribution, the time series data are not always linearly correlated, which hinder the improvement of forecasting efforts. Nevertheless, temperature shows to be the most important climate feature, while precipitation had less influence. As previously stated, the availability of human water sources appears to have a greater impact on the breeding of invasive *Aedes* mosquitoes than natural rainfall, as these mosquitoes often rely on artificial containers near human habitats [34]. Although heavy rainfall can disrupt larval development by washing out breeding sites, the connection between precipitation and mosquito populations varies depending on local climate conditions [34].

Additionally, the inclusion of egg abundance proved to be a key predictor. Our findings confirm that incorporating mosquito-related data improves the fitting and forecasting of predictive models. Consequently, continuous monitoring of mosquitoes and egg abundance by public health systems is essential for more accurate forecasting and effective control measures.

Furthermore, selecting the appropriate lagged variables and ovitrap egg counts, we validated the models using different evaluation metrics. Based on metrics such as Root Mean Squared Error (RMSE) and Mean Absolute Error (MAE), the Random Forest (RF) model outperformed the others, followed by the Seasonal Autoregressive Integrated Moving Average with Exogenous variables (SARIMAX) model. Among the models evaluated, RF performed best on the training data, while the Generalized Linear Model (GLM) performed best on the testing data, with SARIMAX in second place.

The poor performance of the models on the Bizkaia test set can be attributed to differences in the characteristics of the training and test data. Notably, the mean value of the training data is significantly lower than that of the test data, with egg counts in 2023 being unusually high. This discrepancy between the training and test datasets likely contributed to the models’ suboptimal performance for Bizkaia. Nevertheless, for predicting egg abundance in the municipality of Bilbao (Bizkaia), SARIMAX demonstrated superior performance.

Finally, we applied the best-performing models to estimate *Aedes* invasive mosquito abundance in the Basque Country provinces for the upcoming year. By analyzing mosquito egg counts and environmental factors, this study improves and contributes the understanding of seasonal influences on mosquito abundance in a non-endemic region with a maritime climate, characterized by cooler temperatures, rainy weather, and the presence of competent mosquito vectors. These predictions could be used to inform public health strategies and mosquito control efforts, thereby helping to prevent the spread of mosquito-borne diseases in non-endemic regions.

These findings provide valuable insights for future research on assessing the risk of arbovirosis outbreaks in non-endemic regions like the Basque Country. By considering factors such as imported cases, mosquito abundance, and seasonal variations, risk evaluations for mosquito-borne diseases can be refined. Nevertheless, limitations remain in generalizing these results across the diverse areas within each province. For example, Bizkaia, which houses the largest human population in the Basque Country, includes regions with distinct micro-climates that may influence invasive mosquito abundance differently.

Moreover, by considering shorter temporal intervals, such as weekly data collection (depending on vector monitoring schedules and data availability), would improve the precision of vector control strategies and strengthen the assessment of mosquito-borne disease risks. However, this would depend mostly on the vector population monitoring intervals and the availability of data. Furthermore, future improvements in this research should consider a deeper analysis of the methods for partitioning the dataset into training and testing sets, which might enhance the model’s performance.

This research aims to offer an estimate of mosquito population abundance and contribute to the development of vector control strategies, thus mitigating the risks of mosquito-borne infections, particularly considering the region’s specific environmental conditions. Furthermore, this study highlights the critical need for ongoing, localized surveillance to better understand and address the expanding threat of mosquito-borne diseases.

## Declarations

## Acknowledgments

We thank NEIKER, the Basque Institute for Agricultural Research and Development, the Public Health Epidemiological Unit, and the municipalities direction of Basque Country for providing the mosquito eggs count monitoring data. Hamna Mariyam K.B. acknowledge the School of Data Analytics Mahatma Gandhi University in Kottayam, Kerala, India for the support.

## Ethical approval

Not applicable.

## Data availability

The environmental data used in this study were retrieved from several meteorological stations managed by Euskalmet, the Basque Agency of Meteorology. This data is openly available through the OpenData Euskadi platform [29].

The mosquito egg counts, collected using ovitraps, were provided by NEIKER, the Basque Institute for Agricultural Research and Development (for details, see [9]). Due to ethical considerations and commercial sensitivity, these data are not publicly available.

## Supplementary Material

### A. Dataset summary

#### A.1 Climatic variables per province

Figure 10 shows the distribution of climatic data, including temperature, humidity, and precipitation, across the provinces of Gipuzkoa, Bizkaia, and Araba. The graphs summarize the weather variables, highlighting outliers (represented as single points or circular dots) in the dataset. The horizontal line dividing the box in two represents the median value of the time series for each climatic variable.

**Figure 10:**
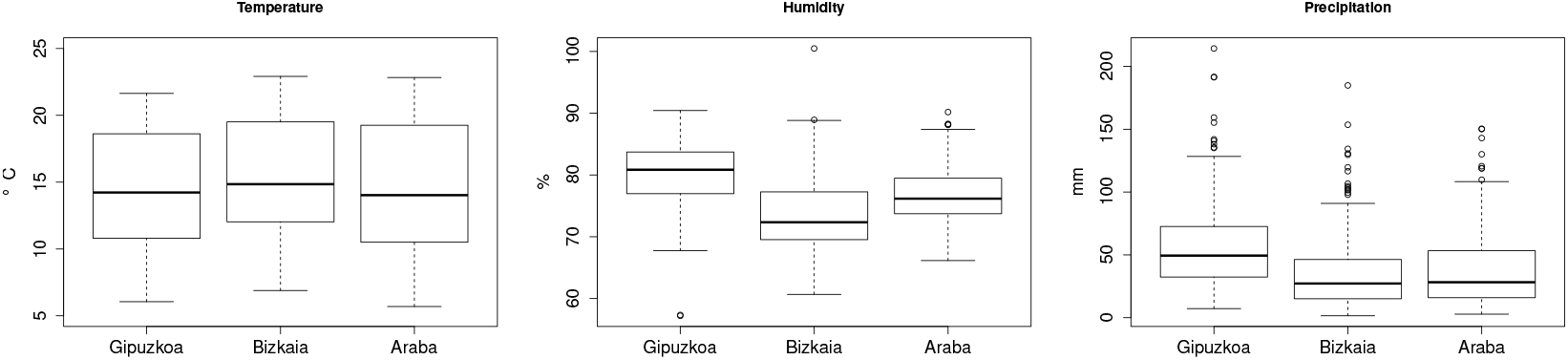
Distribution of time series values for average temperature (°C), relative air humidity (%), and cumulative precipitation (mm) over an interval of 14 days, for all three provinces of the Basque Country.

#### A.2 Lagged time series for Gipuzkoa

Figure 11 (a) shows the monotonic correlation, using Spearman correlation, between the climatic variables and the egg count time series for Gipuzkoa. Figure 11 (b) highlights the time lag at which the highest correlation occurs. At this time lag, the lagged time series will be used as predictors.

**Figure 11:**
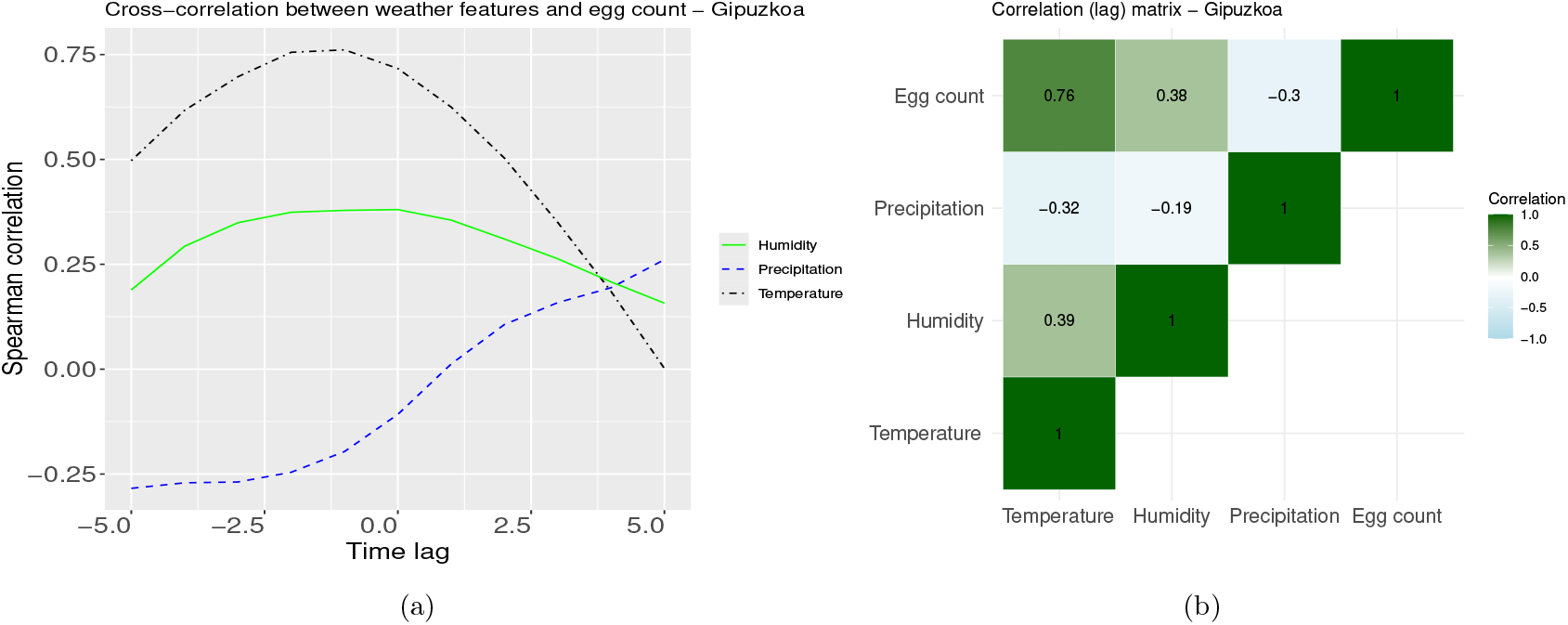
(a) Spearman correlation indices for the time lag between temperature, humidity, precipitation, and the number of eggs. (b) The highest Spearman correlation values between the lagged time series.

#### A.3 Lagged time series for Bizkaia

Figure 12 (a) shows the monotonic correlation using Spearman correlation between the climatic variables and the egg count time series for Gipuzkoa. Figure 12 (b) highlights the time lag at which the highest correlation value occurs. The lagged time series corresponding to this highest correlation will be used as predictor variables.

**Figure 12:**
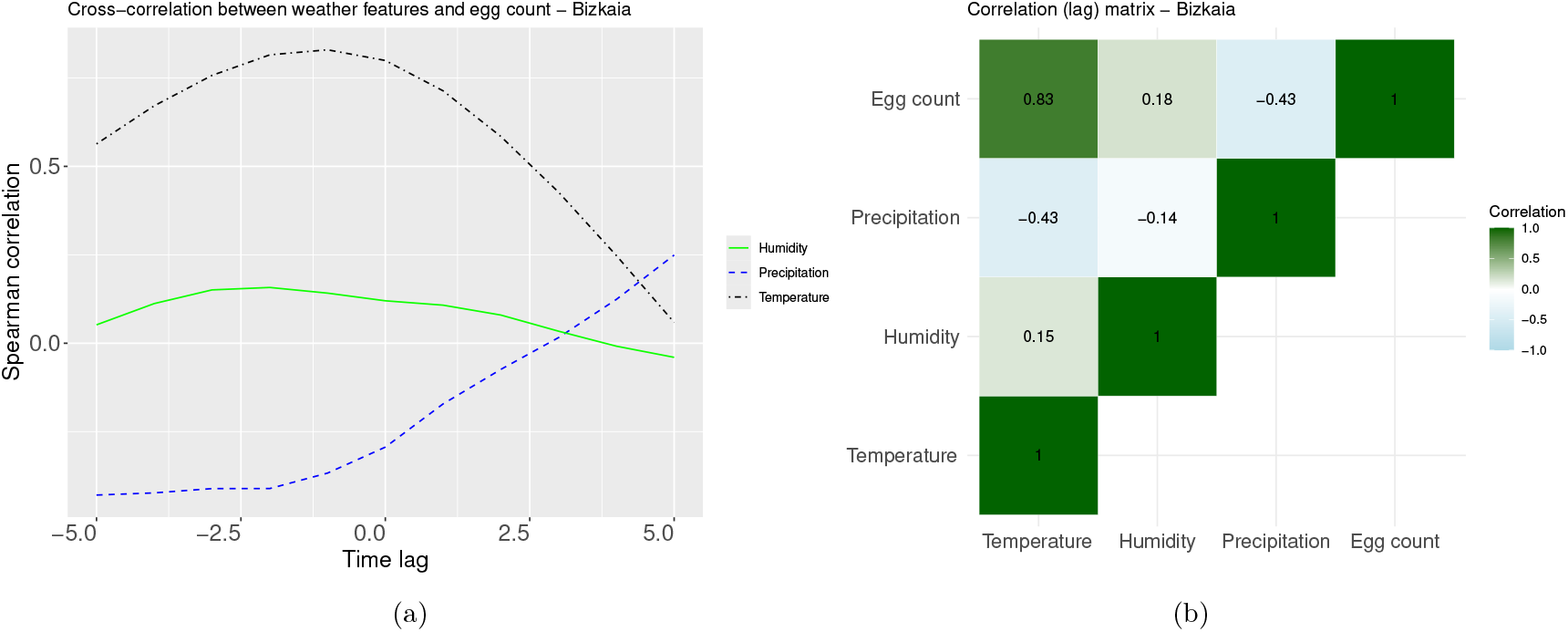
(a) Spearman correlation indices for time lags between temperature, humidity, and precipitation with the number of eggs. (b) The highest Spearman correlation values for the lagged time series.

### B. Data selection per municipalities

Due to the dispersed data with many zero values for egg counts and the lack of sufficient information to create a reliable training dataset at the municipality level, we conducted the analysis at the provincial level. However, we selected one municipality from each province to present the results.

For Gipuzkoa, we selected Irun, a municipality of interest due to its proximity to the French border and the frequent movement of travelers. Irun also had more positive ovitraps during the analysis period compared to the capital, Donostia/San Sebastián. The C084 weather station in Irun was selected over C083, as the latter’s dataset lacked temperature, precipitation, and humidity data.

For Bizkaia, we chose Bilbao as the municipality, rather than larger municipalities like Barakaldo or Basauri, since both lack meteorological stations within their boundaries. For Bilbao, station C0B0 had no data on precipitation or temperature, while station C039, located in Deusto, provided data from 2016 to 2021, and station C03A began recording data in December 2021. To cover the entire study period, data from both C039 and C03A were used in the initial analysis.

For Araba, we chose the municipality of Laudio/Llodio. The primary reason for selecting this municipality is that no egg counts were recorded in the ovitraps in the capital, Vitoria. For weather data, two meteorological stations in Laudio/Llodio were listed in the database (see more details in [29]): station C067, which was selected, and station C027, which was not included due to missing data for the chosen period.

Egg count data were collected at the municipality level, disregarding specific ovitrap locations. The dataset was constructed by averaging the highest three egg counts from the ovitraps for each municipality every 14 days (bi-weekly). This approach was necessary because the number of monitored ovitraps varied throughout the study period. We selected the top three counts since, on average, no more than 10 ovitraps were placed in each municipality every 14 days. Weather data were aggregated by municipality on a daily basis. A dataset was then constructed containing the average temperature, average humidity, and cumulative precipitation for the previous 14 days.

This information was combined into a single dataset for each municipality. The time series of egg counts, average temperature, humidity, and cumulative precipitation are presented in Figures 13(a)-(b), 13(c)-(d), and 13(e)-(f).

**Figure 13:**
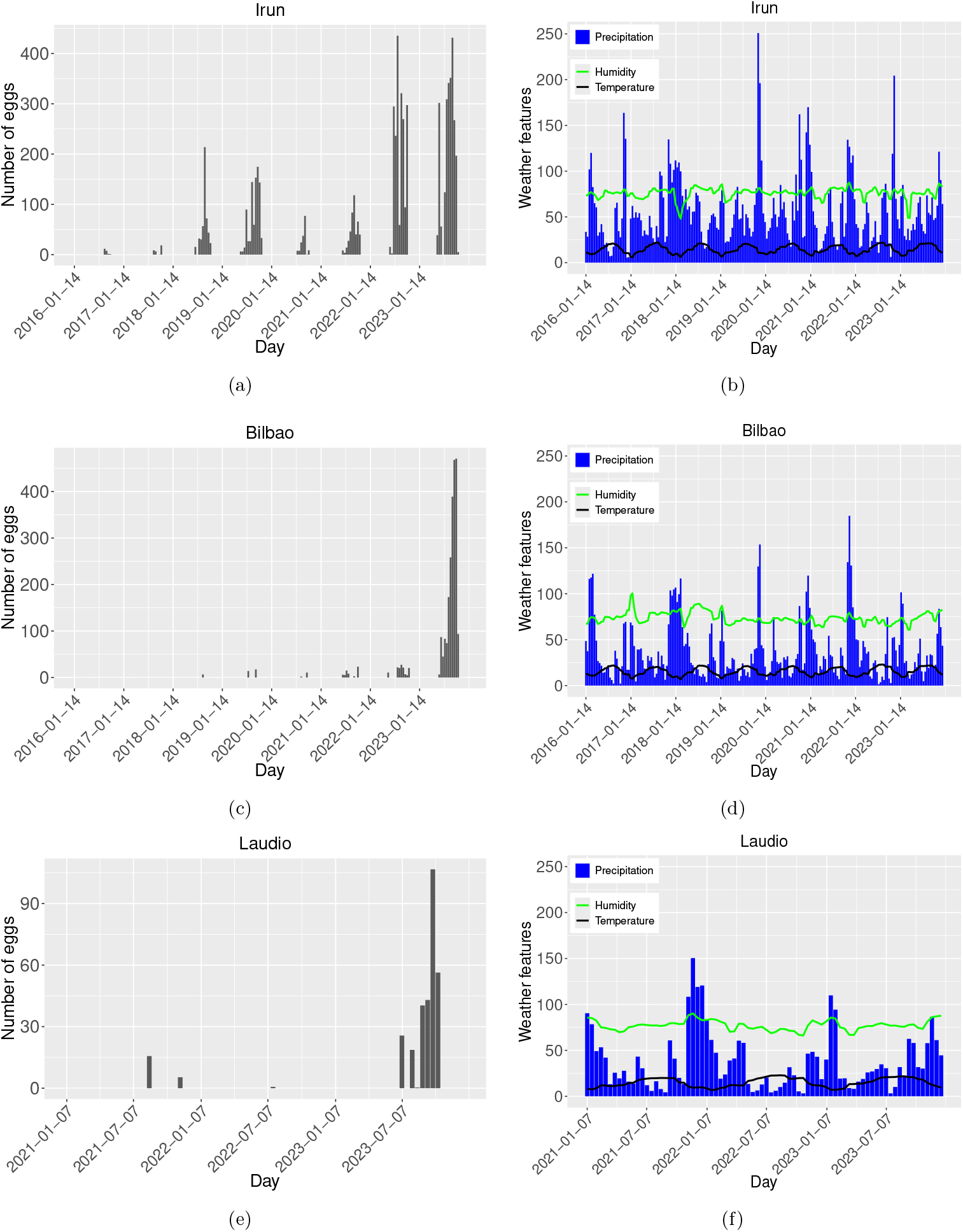
Number of mosquito eggs collected, in (a), (c), (f), and average temperature (°*C*), relative air humidity (%), and cumulative precipitation (*mm*), in (b), (d), (f). For Irun (in Gipuzkoa), Bilbao (in Bizkaia) and Lodio (in Araba).

For further analysis, we will focus on the municipalities of Irun and Bilbao, since Laudio has only recorded positive ovitraps from 2021 onward. The same methodology and analysis applied at the provincial scale will now be carried out at the municipality scale.

#### B.1 Irun

##### Statistical analysis

For Irun, Figure 14 shows the relationship between meteorological variables and number of mosquito eggs count using scatter plots. While Figure 15 shows the monotonic correlation using Spearman correlation between eggs counts and the time lagged versions of the climate features. For temperature, maximum correlation occurs at -1 units (2 weeks). For humidity, maximum occurs at -2 units. And for precipitation, maximum correlation occurs at -5 units, with negative correlation.

**Figure 14:**
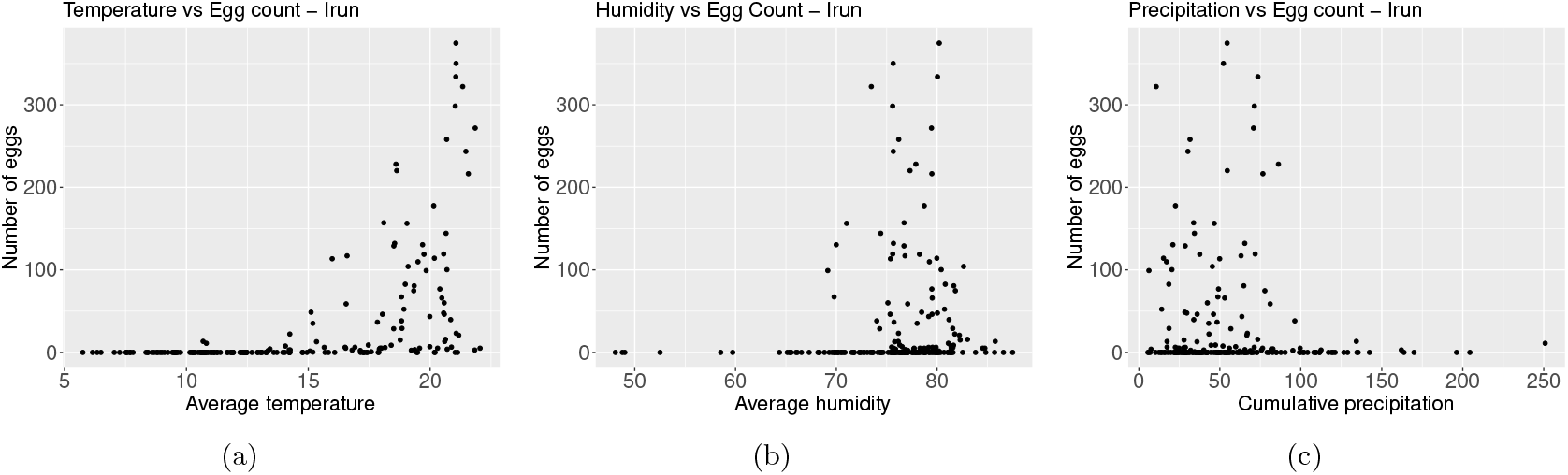
(a) Average temperature (in °*C*) versus number of collected mosquitoes eggs. As temperature increases, the number of eggs increases. (b) Average air relative humidity (in %) versus number of collected mosquitoes eggs. As humidity increases, the number of eggs increases. (c) Accumulated precipitation (in *mm*) versus number of collected mosquitoes eggs. As precipitation increases, the number of eggs decreases.

**Figure 15:**
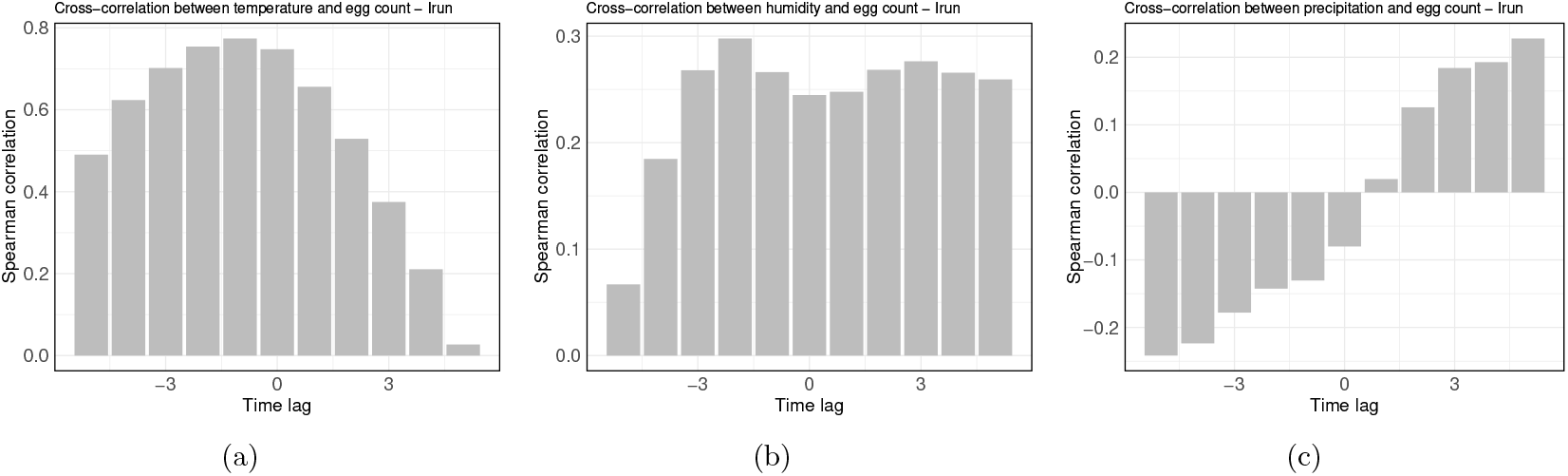
Spearman correlation between the weather feature time series and the number of mosquito eggs, with a time lag of 1 unit (2 weeks). (a) For temperature, the maximum correlation occurs at a lag of −1 unit. (b) For humidity, the maximum correlation occurs at a lag of −2 units. (c) For precipitation, the maximum negative correlation occurs at a lag of −5 units.

##### Fitting

We implemented the GLMG, SARIMAX, RF, and CT models using the R programming language for the Irun dataset. The training dataset spans from 2017 to 2022, while the test dataset consists of data from 2023, as shown in Figure 16. We compared the models’ performance on both the training and testing datasets, evaluating them using the Mean Absolute Error (MAE), Root Mean Squared Error (RMSE), and R-squared score (R^2^), as detailed in Table 1.

**Table 1:** Evaluation of error metrics for each model on the training and testing datasets (for Irun). MAE represents the Mean Absolute Error, RMSE the Root Mean Squared Error, and *R*^2^ the R-squared score.

| Model | Evaluation Metrics |  |  |  |  |  |
| --- | --- | --- | --- | --- | --- | --- |
| | MAE Train | MAE Test | RMSE Train | RMSE Test | $R^2$ Train | $R^2$ Test |
| RF | 4.00 | 43.27 | 10.22 | 68.84 | 0.97 | 0.70 |
| SARIMAX | 8.45 | 37.55 | 17.32 | 57.35 | 0.91 | 0.79 |
| GLM | 10.10 | 33.51 | 21.57 | 52.87 | 0.86 | 0.82 |
| CT | 11.77 | 54.21 | 25.86 | 89.16 | 0.80 | 0.49 |

**Figure 16:**
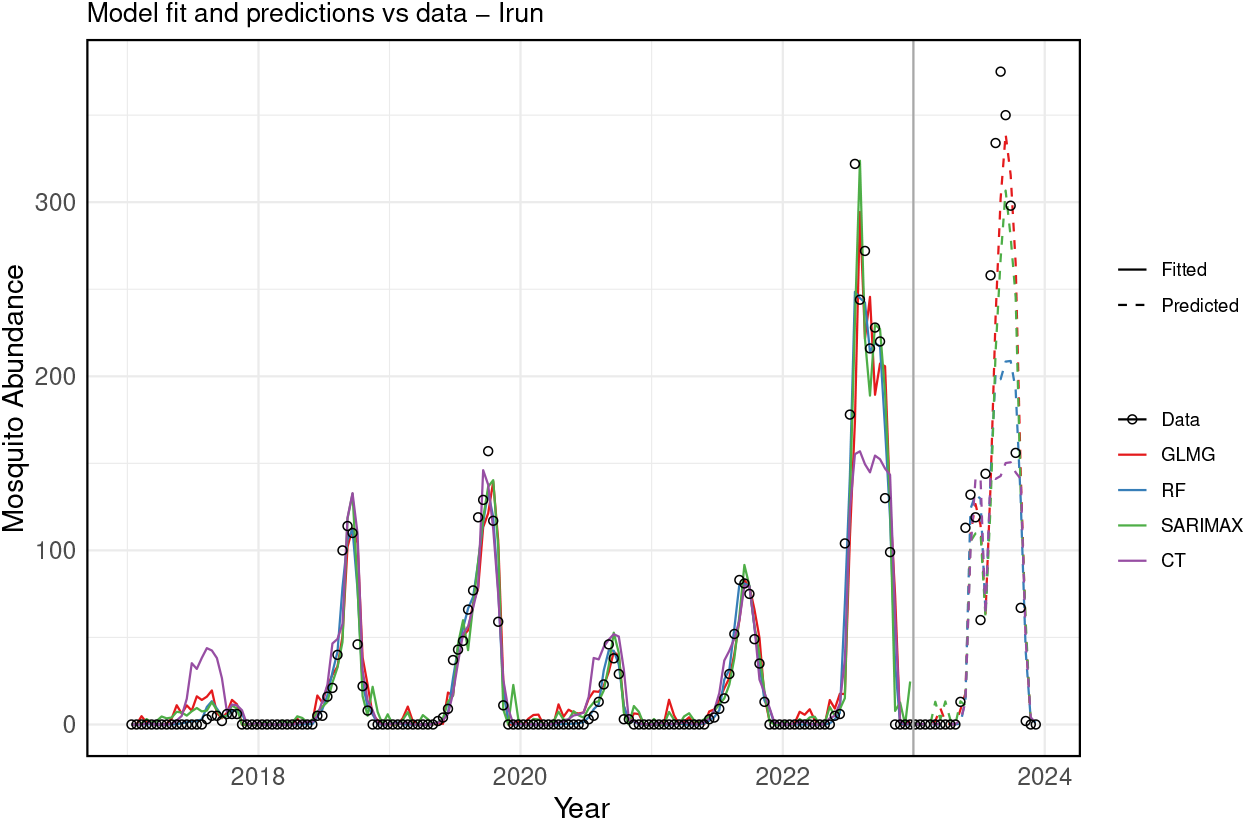
Comparison of actual data versus fitted and predicted values for the Irun dataset. The actual data is represented by open black circles, while the fitted values for each model are shown as solid lines and the predicted (test) values as dashed lines. The models are colored as follows: RF model (`ntree` = 600, `mtry`= 5) in blue, GLMG model in red, SARIMAX model in green, and CT model (`ntree` = 500, `mtry` = 3) in purple. The vertical gray line separates the training dataset (2017–2022) from the testing dataset (2023).

##### Forecasting

Subsequently, we used the best-performing trained models to forecast future *Aedes* invasive mosquito abundance in Irun. To do this, we included 2023 data points as part of the training dataset. Using the historical time series data and their lagged versions, we predicted future values based on the most recent observations (see Figure 17). The error analysis for the training dataset is presented in Table 2, which shows that the Random Forest (RF) model performed the best, explaining 97% of the variance in the training dataset.

**Table 2:** Evaluation of error metrics in the training dataset (for Irun), highlighting the best model performance. MAE refers to the Mean Absolute Error, RMSE stands for the Root Mean Squared Error, and *R*^2^ represents the R-squared score.

| Evaluation Metrics in the training dataset |  |  |  |
| --- | --- | --- | --- |
| Model | MAE | RMSE | $R^2$ |
| RF | 5.90 | 12.17 | 0.97 |
| SARIMAX | 11.89 | 23.56 | 0.90 |
| GLM | 13.68 | 27.71 | 0.86 |

**Figure 17:**
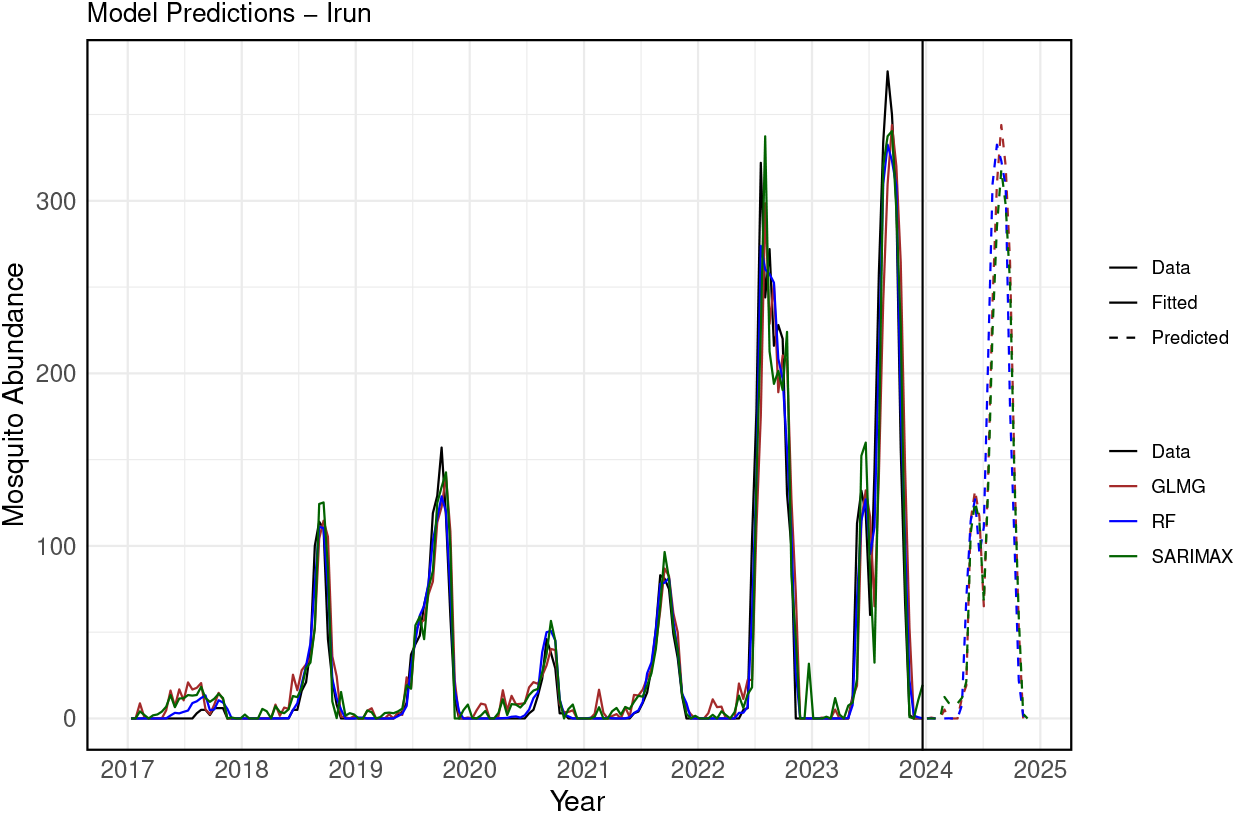
The actual data versus the fitted and predicted values of the model for Irun. The data is represented by a solid black line, with fitted values shown as solid lines in color and predicted values as dashed lines. In blue, the RF model (`ntree` = 600, `mtry` = 5); in brown, the GLMG model; and in green, the SARIMAX model. The vertical black line separates the training dataset (from 2017 to 2023) from the forecasted period for the year 2024.

#### B.1. Bilbao

##### Statistical Analysis

For Bilbao, Figure 18 presents the relationship between meteorological variables and mosquito egg counts using scatter plots. Figure 19 illustrates the monotonic correlation, calculated using Spearman’s correlation, between egg counts and the time-lagged versions of the climate features.

**Figure 18:**
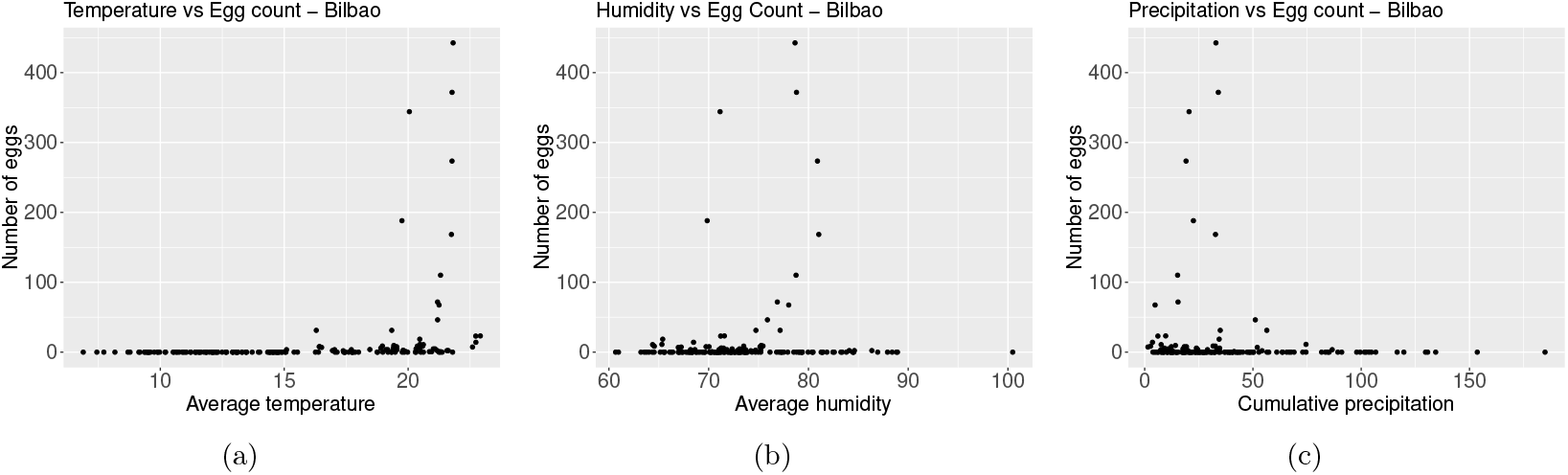
(a) Average temperature (in °*C*) versus number of collected mosquito eggs. As temperature increases, the number of eggs increases. (b) Average relative humidity (in %) versus number of collected mosquito eggs. As humidity increases, the number of eggs increases. (c) Accumulated precipitation (in *mm*) versus number of collected mosquito eggs. As precipitation increases, the number of eggs decreases.

**Figure 19:**
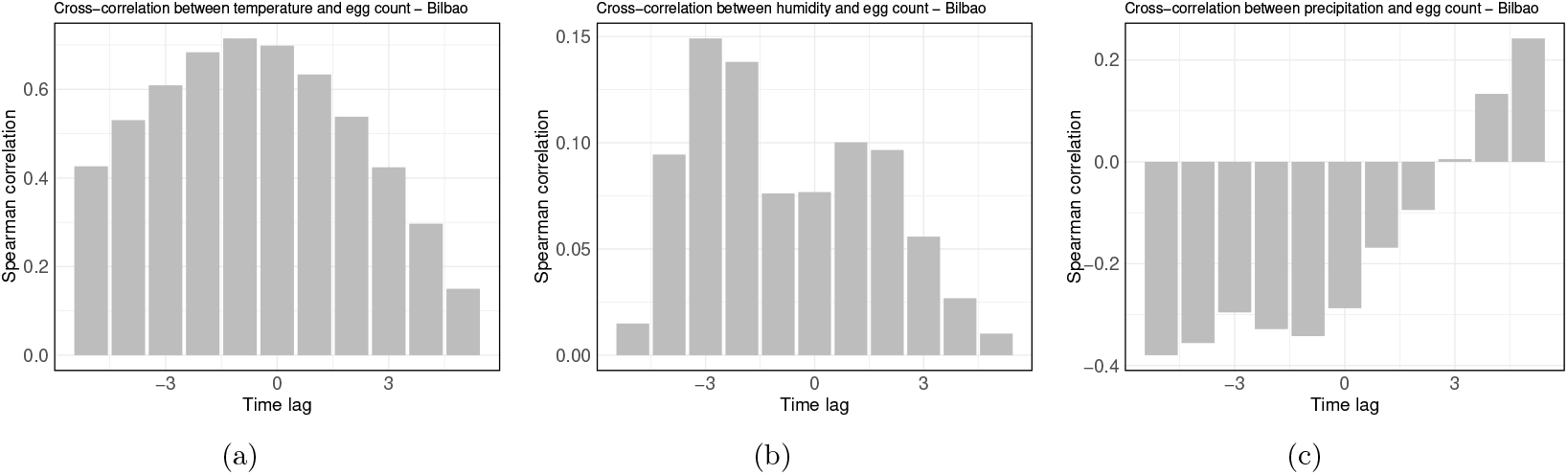
Spearman correlation between the weather features time series and number of mosquito eggs with a time lag of 1 unit (2 weeks). For (a) temperature, maximum correlation occurs at −1 units. For (b) humidity, maximum correlation occurs at −3 units. For (c) precipitation, maximum correlation occurs at −5 units.

##### Fitting

We also implemented the GLMG, SARIMAX, RF, and CT models for the Bilbao dataset. The training dataset consists of data from the year 2019 to 2022, while the test dataset consists of data points from 2023, as shown in Figure 20. We compare the models on both the training and testing datasets, evaluating each model’s performance using the MAE, RMSE, and R^2^ metrics, as detailed in Table 3.

**Table 3:** Different evaluation error metrics in the train and in the test dataset (for Bilbao) of each model chosen. MAE for the Mean Absolute Error, RMSE for the Root Mean Squared Error, and *R*^2^ representing the R-squared score.

| Model | Evaluation Metrics |  |  |  |  |  |
| --- | --- | --- | --- | --- | --- | --- |
| | MAE Train | MAE Test | RMSE Train | RMSE Test | $R^2$ Train | $R^2$ Test |
| RF | 0.57 | 81.38 | 0.99 | 150.86 | 0.95 | -0.32 |
| SARIMAX | 1.10 | 45.02 | 1.75 | 81.82 | 0.86 | 0.61 |
| GLM | 1.25 | 36.78 | 2.25 | 64.42 | 0.76 | 0.76 |
| CT | 1.41 | 82.00 | 2.72 | 152.01 | 0.65 | -0.34 |

**Figure 20:**
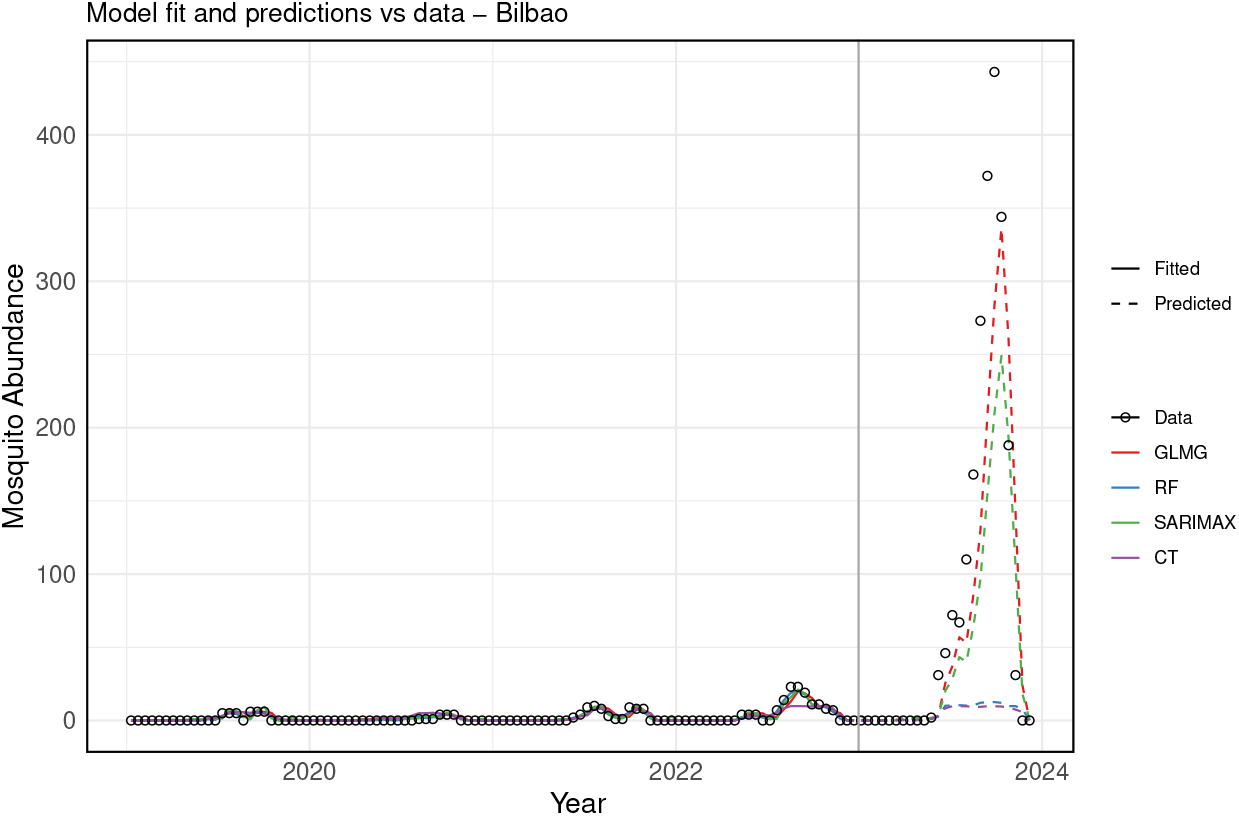
The actual data versus the fitted a nd tested values o f the m odel f or I run. The d ata is represented by open black circles, while the fitted values of each model are shown in solid lines and the predicted (tested) values in dashed lines. In blue, the RF model (`ntree`= 600, `mtry` = 5); in red, the GLMG model; in green, the SARIMAX model; and in purple, the CT model (`ntree` = 500, `mtry` = 3). The vertical gray line delineates the training dataset (from 2017 to 2022) from the testing dataset (2023).

##### Forecasting

The best-trained model was then used to predict future *Aedes* invasive mosquito abundance in Bilbao using the historical time series data and lagged versions, as shown in Figure 21. The error analysis for the training dataset is presented in Table 4, indicating that the model with the best performance is the SARIMAX model, which explains 97% of the variability in the training dataset.

**Table 4:** Evaluation error metrics for the training dataset (for Bilbao), showing the best model performance. MAE represents the Mean Absolute Error, RMSE is the Root Mean Squared Error, and *R*^2^ denotes the R-squared score.

| Evaluation Metrics in the training dataset |  |  |  |
| --- | --- | --- | --- |
| Model | MAE | RMSE | $R^2$ |
| RF | 4.98 | 15.28 | 0.95 |
| SARIMAX | 4.63 | 10.57 | 0.97 |
| GLM | 9.45 | 24.23 | 0.87 |

**Figure 21:**
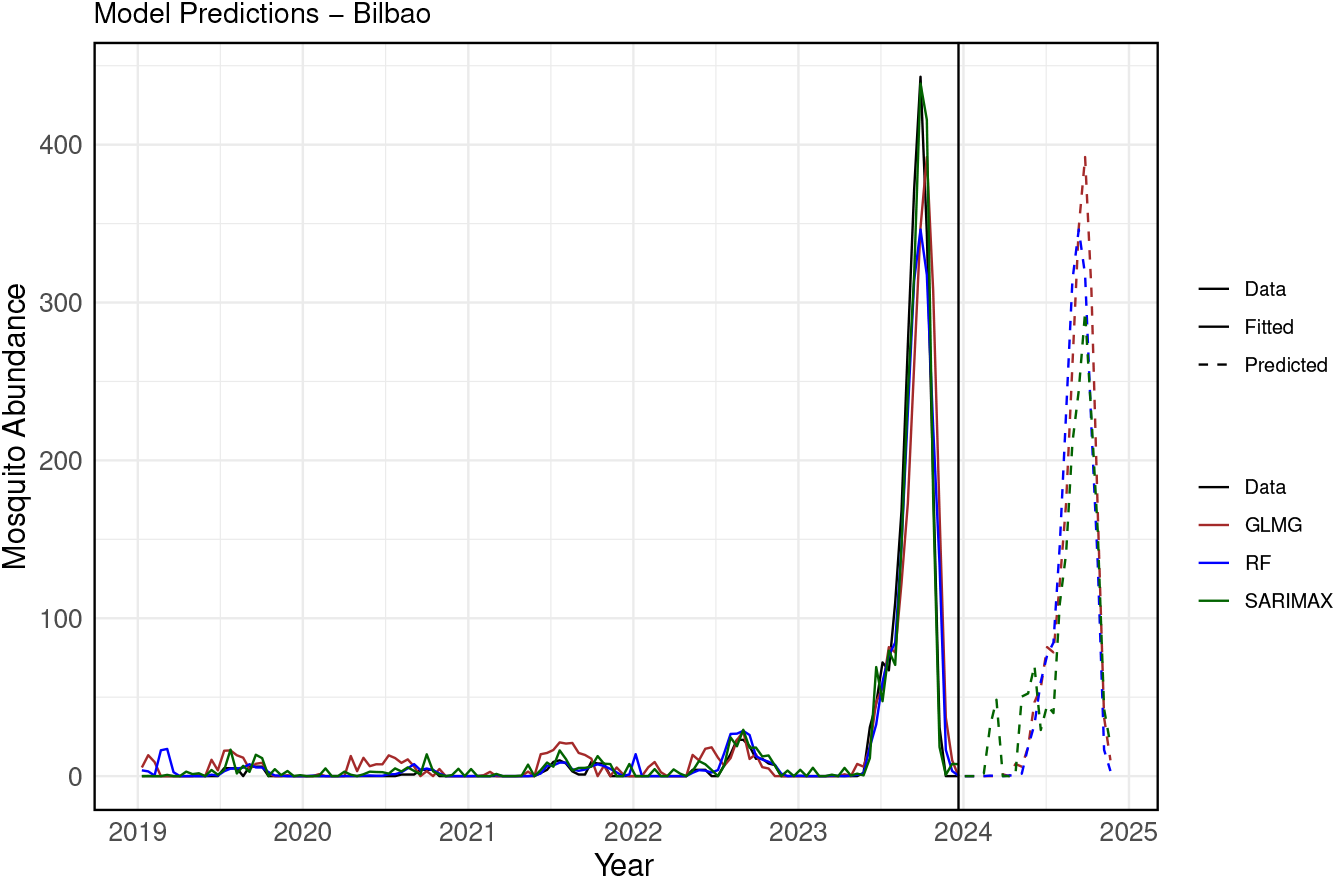
Actual data versus fitted and predicted values for Bilbao. The actual data is represented by a solid black line. The fitted values for each model are shown in solid colored lines, while predicted values are in dashed lines. The RF model is shown in blue (`ntree` = 600, `mtry` = 5), the GLMG model in brown, and the SARIMAX model in green. The vertical black line separates the training dataset (from 2017 to 2023) from the forecasted period for the year 2024.

## Footnotes

1 Note that not all meteorological stations displayed on the map contain records of all environmental features selected for this study.

